# Data Preparation of the nuMoM2b Dataset

**DOI:** 10.1101/2021.08.24.21262142

**Authors:** Anton Goretsky, Anastasia Dmitrienko, Irene Tang, Nicolae Lari, Owen Kunhardt, Raiyan Rashid Khan, Cassandra Marcussen, Adam Catto, Daniel Mallia, Alisa Leshchenko, Adam (Yun Chao) Lin, Anita Raja, Ansaf Salleb-Aouissi, Itsik Pe’er, Ronald Wapner, Cynthia Gyamfi-Bannerman

## Abstract

In 2010, the Eunice Kennedy Shriver National Institute of Child Health and Human Development (NICHD) started the Nulliparous Pregnancy Outcomes Study: Monitoring Mothers-to-be (nuMoM2b), a prospective cohort study of a racially/ethnically/geographically diverse population of nulliparous women with singleton gestation. The nuMoM2b is a very large dataset, consisting of data for 10,038 patients with over 4,600 features per patient, spread out over 80 files. In this report, we share our experience preparing and working with this dataset. We present our data preprocessing of the nuMoM2b dataset to get a deeper understanding of the data, extract the most relevant features, make the fewest assumptions when filling in unknown values, and reducing the dimensionality of the data. We hope this report is useful to researchers interested in building machine learning and statistical models from the nuMoM2b dataset.

## 1 Introduction to nuMoM2b and Data Processing

The primary goal of the nuMoM2b study [1] was to determine the maternal characteristics, both clinical and genetic factors, physiological response to pregnancy and environmental factors that could be used to derive models that accurately predict adverse pregnancy outcomes (APOs). Our team has extensive prior experience working on medical data for preterm birth (PTB) prediction [2, 4, 5].

As originally organized, the dataset is not immediately conducive to analysis to those unfamiliar with the medical background, nor is it conducive to quick placement into machine learning models. The ratio of instances to features would result in an inevitable model overfitting. Various medical categories exist within an individual file, data relevant to features exist throughout many different files, and dependencies and redundancies exist across the whole dataset. As such, nuMoM2b required extensive review and processing of features, their dependencies and relations in order to reduce the complexity of the dataset, and shape it in a form amenable to a variety of machine learning (ML) algorithms and exploratory data analysis (EDA).

The intended audience for this document is researchers interested in building machine learning and statistical models from the nuMoM2b dataset. Our aim is to share our experience preparing and working with this dataset. The intent is not to share the processed dataset nor the scripts that are specific to research aims.

Our specific research goal is to build machine learning models for the prediction and prevention of preterm birth in nulliparous women using the nuMoM2b dataset. This project is funded by the NIH/NLM (Project # 1R01LM013327-01). The data review and processing work described in this document was conducted through the direct collaboration of the Computer Science departments at Hunter College and Columbia University, along with maternal and fetal medicine experts at the Columbia University Medical Center. The following are the goals of this collaborative effort:

- Significant reduction of the feature space for a better management of the data and a a reduction of the risk for overfitting.
- A reformatting of the dataset, to allow for easy configuration of the data, conducive to exploration and machine learning modeling.
- Extensive literature review of existing risk factors related to data categories and causal pathways of PTB.
- Exploratory data analysis of both the reduced and the unmodified feature space.

As a result of the effort, the dataset was reduced to 364 features at the most general level of complexity, and 465 features at a higher level of detail. Extensive filtering and imputation rules were created to accomplish this goal, along with a system for both human readability and easy script interpretation. An extensive literature review was performed documenting odds ratios. EDA was performed on the dataset, comparing calculated odds ratios to the literature review, and discovering and correcting data inconsistencies. Finally, we summarize our thoughts on data preprocessing and nuMoM2b in Section 7.

We summarize and visualize the PTB statistics in the dataset in Figure 1 and 2. See [3] for more details about the dataset.

**Figure 1:**
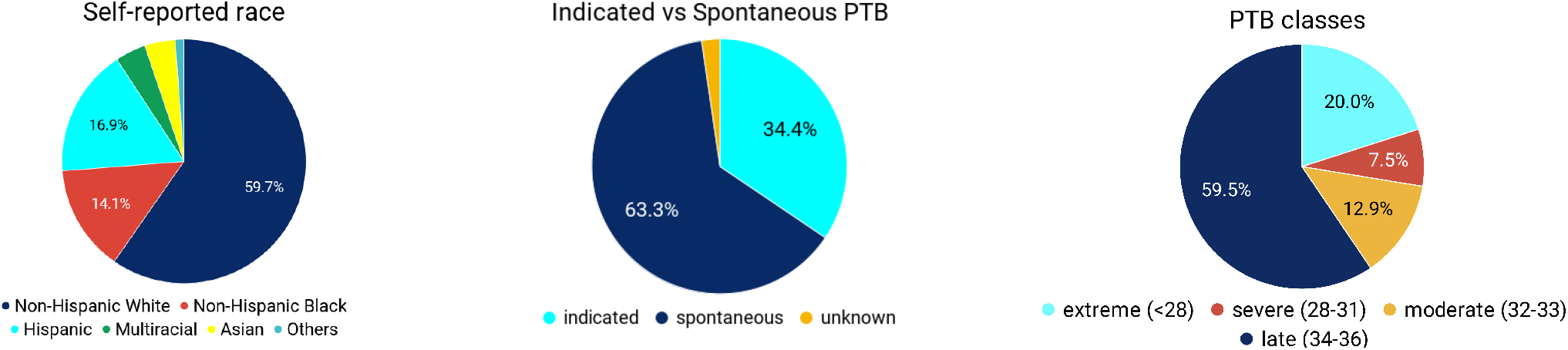
Preliminary statistics in the nuMoM2b data

**Figure 2:**
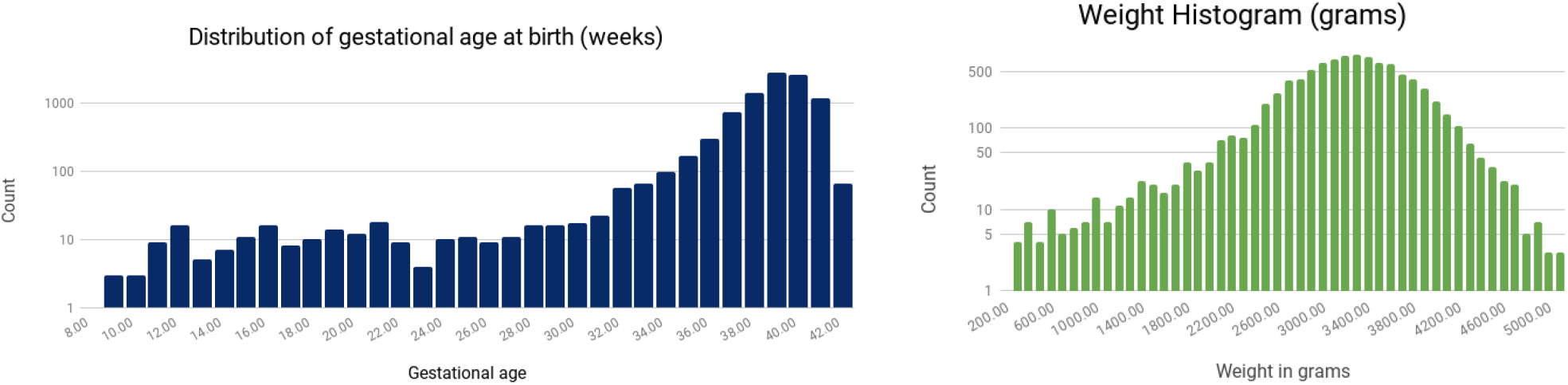
Distribution of gestational age and weight in the nuMoM2b data

## 2 Preprocessing

As part of the preprocessing stage, the information present in the provided codebooks on basic variable statistics, distribution, and type was transferred over via a script to a workable spreadsheet. Then to support our analysis, the following steps were taken:

- **“Data type” was corrected to a more accurate label**. Many variables were labeled “num” for numeric even though they actually represented categorical data. For example, Country of Birth (V1AF08) was labeled numeric. However, country cannot be treated as a numeric feature in modeling. Numeric, according to the codebook, simply meant the values used in labeling were numerals. This problem was repeated across the entire dataset. As such, each variable was looked at manually and categorized properly for typical modeling and analysis. Many variables labeled “num” were changed to “categorical”.
- **A “Missingness” metric was calculated** using the variable statistics provided in the codebook to help with data analysis.
- **A new “Temporal” label was added** in order to label variables that appear at multiple unique time points throughout the study. nuMoM2b data was recorded at several distinct time points, and some variables are updated over time / questions are asked more than once. Temporal had values of True or False.
- **A new “Temporal Detail” label was added** in order to label at which time point a variable is relevant. Not all data collected are relevant only to the moment of collection in the study. For example, many demographic related questions were asked at Visit 2. However, those are static features and significant at all points in the study. This concept will be explained further under the section titled “Timeline”.

Accuracy of action timing, such as treatment administration like medications, and the creation of these “temporal” labels are important for this team’s goals of sequential treatment decision making modeling.

## 3 Organization into Groups

After the preprocessing stage, further organization was desired to better understand and manage the dataset. In order to better organize the data, **Filtering Groups** were created. The goal of these groups was to break down the dataset into more understandable portions for those unfamiliar with the detailed medical content, and for simpler processing of data in bulk. The following were the filtering groups created for the nuMoM2b dataset.

- **Treatment** - Variables relating to intervention in cases of potential or immediate PTB risk, such as progesterone administration, steroids, and last minute medical administrations at delivery.
- **Psychological** - Variables relating to the psychological state of the patient, through multiple scales.
- **Physiological** - Variables relating to instantaneous measurements of physiology, such as temperature, symptoms of flu-like illnesses, blood pressure, etc.
- **Medical History** - Variables relating to long term medical history and conditions, but also various tests performed on both the mother and the fetus or newborn found in the study.
- **Demographics** - Variables relating to the patient’s demographic factors, such as race, income, education, etc.
- **Ultrasound** - Variables that were recorded from the research and clinical ultrasounds the patient went through and marked as such in the dataset.
- **Outcomes** - A metadata file containing various variables that are useful mainly as classifier labels, or are features that were collected post-delivery about the mother or newborn.
- **Activity** - Variables relating to the physical activity of the patient.
- **Toxicology** - Variables relating to the medications taken just before and during pregnancy, and / or their relations to particular reasons / conditions.
- **Family History** - Variables relating to family medical conditions and history.
- **Food Frequency Analysis** - Variables relating to food, diet and vitamins, in the three months prior to pregnancy.
- **Sleep Substudy** - Select variables from the two sleep substudies included in nuMoM2b.

## 4 Processing

Now that the dataset was more simply organized, the process of feature reduction began. Filtering groups were divided among teams, who worked in consultation with the Columbia University Medical Center OB/GYN collaborators, to determine which features to keep as is, which features to summarize into scales, scores, or other aggregate forms, and which features to remove for redundancy or other reasons. As a result of this effort, a system was created to organize the efforts in a form both readable by people, and interpretable by scripts, to allow for easy updates. This system consists of the creation of **Filtering** and **Imputation Rules**, and **Layering**. Filtering rules serve to keep, drop, or summarize features, while imputation rules serve to impute missing data as required by many modeling algorithms. Layering served to organize the data into different levels of abstraction from the most general to the most specific level of information.

### 4.1 Layers

Layers were decided upon given the high feature complexity of nuMoM2b, even after much feature reduction. As many questions are structured around a format of a general question followed by several sub-questions, it was reasoned that the general question should in most cases be representative of the data points that follow. For example, V1AD06 asks, *Have you had any ‘flu-like illnesses’, ‘really bad colds’, fever, a rash, or any muscle or joint aches since you became pregnant?* This question is then followed by questions in regard to which symptoms are actually present in this ‘flu-like illness’. V1AD06 covers all, and is such a more general question, and thus would be selected into a more general layer, while symptom specific questions would be reserved for the detail-oriented layers, or dropped. If dropped, they may be brought back if desired, or if some significance is found in the most general feature. Internally, we decided upon 3 layers.

- Layer 0 would consist of known risk factors for PTB, along with variables shown to have high odds ratios in our EDA.
- Layer 1 would be the most general layer, consisting of L0 and all general questions that cover as much information as possible.
- Layer 2 would bring back detail that may have been lost, or not included given the generalization and simplicity of L0 and L1.

We will not go into variable-level detail in each layer, but we believe this concept can serve as an organizational method for large complex datasets.

### 4.2 Filtering and Imputation Rules

As part of the data cleaning process, data first passes through a general filtering script, with the rules shown in Table 1. It then passes through an imputation script, with the rules shown in Table 2. Throughout the processing of this dataset, we strived to hold to a set of generally applicable rules for imputation. Below is a sample of the imputation rules used for nuMoM2b.

**Table 1:**
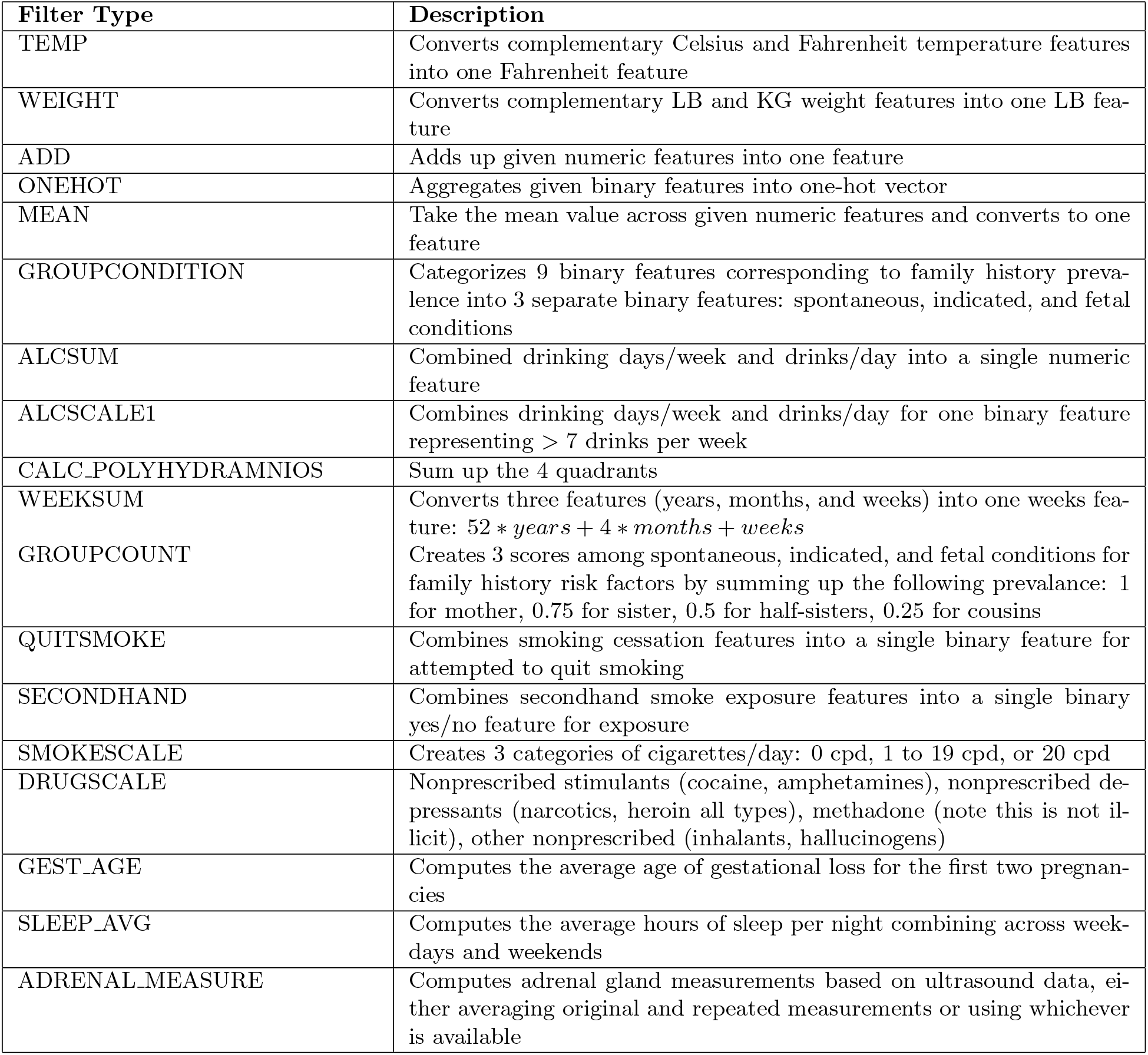
List of Filter Rules and their Descriptions

| Filter Type | Description |
| --- | --- |
| TEMP | Converts complementary Celsius and Fahrenheit temperature features into one Fahrenheit feature |
| WEIGHT | Converts complementary LB and KG weight features into one LB feature |
| ADD | Adds up given numeric features into one feature |
| ONEHOT | Aggregates given binary features into one-hot vector |
| MEAN | Take the mean value across given numeric features and converts to one feature |
| GROUPCONDITION | Categorizes 9 binary features corresponding to family history prevalence into 3 separate binary features: spontaneous, indicated, and fetal conditions |
| ALCSUM | Combined drinking days/week and drinks/day into a single numeric feature |
| ALCSCALE1 | Combines drinking days/week and drinks/day for one binary feature representing $> 7$ drinks per week |
| CALC.POLYHYDRAMNIOS | Sum up the 4 quadrants |
| WEEKSUM | Converts three features (years, months, and weeks) into one weeks feature: $52 * years + 4 * months + weeks$ |
| GROUPCOUNT | Creates 3 scores among spontaneous, indicated, and fetal conditions for family history risk factors by summing up the following prevalence: 1 for mother, 0.75 for sister, 0.5 for half-sisters, 0.25 for cousins |
| QUITSMOKE | Combines smoking cessation features into a single binary feature for attempted to quit smoking |
| SECONDHAND | Combines secondhand smoke exposure features into a single binary yes/no feature for exposure |
| SMOKESCALE | Creates 3 categories of cigarettes/day: 0 cpd, 1 to 19 cpd, or 20 cpd |
| DRUGSCALE | Nonprescribed stimulants (cocaine, amphetamines), nonprescribed depressants (narcotics, heroin all types), methadone (note this is not illicit), other nonprescribed (inhalants, hallucinogens) |
| GEST_AGE | Computes the average age of gestational loss for the first two pregnancies |
| SLEEP_AVG | Computes the average hours of sleep per night combining across weekdays and weekends |
| ADRENAL_MEASURE | Computes adrenal gland measurements based on ultrasound data, either averaging original and repeated measurements or using whichever is available |

**Table 2:** List of Imputation Rules and their Descriptions

| Imputation Name | Description |
| --- | --- |
| MEAN | Imputes with the mean (average) of the existing values |
| BMICAT | Imputes based on existing BMI |
| ONEHOT | Encodes current values by assigning each unique value to an index and assigns a vector of zeros for missing values |
| ANY | Imputes with a value of True if any of given features is present: False otherwise |
| MULTI | Imputes with a value of True if any of given features is present: False otherwise (*different internal organization, outcome same as ANY*) |
| SINGLE | Imputes with a value of True if a given feature is present: False otherwise |
| NEG1 | Imputes missing values with -1 |
| NUMERIC | Imputes missing values with the given numeric value |
| MODE | Imputes with the mode of the existing values |
| SUM_LEQ | Imputes with a value of True for Oligohydramnios if sum of Amniotic Fluid Index across 4 quadrants $< 5$: False otherwise |

- If a numeric-like feature is applicable to a vast majority of patients, and the missingness was relatively low, we attempted to impute with a value such as MEAN or MODE. If multiple measurements were made on the same information, the mean took into account all of them (excluding those marked as incomplete or inaccurate).
- If a feature serves as a general precursor to a list of follow-up questions, such as “*Have you had any flu-like illnesses, ‘really bad cold’, fever, a rash*…” followed by questions regarding symptoms, if data is present in the follow-up but the is missing for the general question, we impute the general question to whatever value represents True. Otherwise we often impute to an unknown or not applicable determiner such as 999, especially if assumption is misleading in the understanding of treatment. Often, these imputation rules looked at follow-up questions that were not included in the current layer or were excluded from modeling, due to the general question covering the topic. For the follow-up questions themselves, imputation may have been left at unknown or imputed to a value depending on the missingness and applicability.
- Negative one (−1) was often used to represent unknown or inapplicable for numeric features.
- When there is parent feature and there are several related child features that are too detailed, DROP the child features.

## 5 Timeline

This timeline is meant to serve as a high level representation of the way we processed and organized data for internal analysis and modeling. This timeline does not cover all variables and is thus not an exhaustive representation of nuMoM2b. It serves rather as a quick glance into the data currently in use, accurate at time of publication, but subject to change. Each internal substudy may use a different collection of variables for their goals.

This timeline shows the existence of six unique time points that nuMoM2b represents. 5 of those time points are directly sequential, namely **Before Pregnancy, Visit 1, Visit 2, Visit 3, Visit 4 (Delivery Visit)**, and **Post Delivery**. There also exists a **Constant** time point, which groups data that may have been collected at different points in the study, but applies at all times to the patient, such as race and pregnancy history. In an ideal full-term pregnancy, the patient would pass through all these time points, and have data recorded under the Constant point. However, if a birth was preterm, still, or a patient missed a visit, their timeline could skip some of either Visit 1, 2 or 3, and move straight to Visit 4, which represents data collected at delivery. This is represented by the dashed arrow in the timeline. Variables in the timeline are organized by the Filtering Groups described earlier, and are abstractly summarized and simplified in the tables below. On the bottom right we see two boxed sections. These represent time points that occurred in between time points in this timeline. Antepartum evaluations may have occurred between visits, and enrollment screening occurred before visit 1. ***To reiterate, this timeline is not representative of all data available or used, but should rather serve as a quick guide***.

**Table 3:**
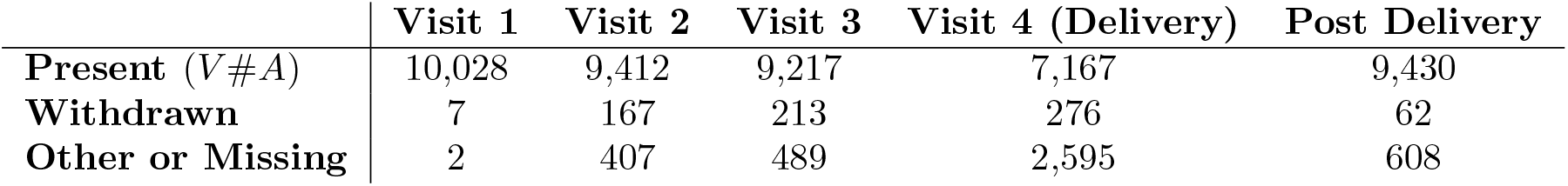
Amount of patients available at each visit. *Present* numbers are patients who filled out the main maternal interview form. Patients designated as other / missing may still have information available at that visit, especially under Chart Abstractions. **Delivery outcomes are available for almost every patient except those withdrawn**. *Withdrawn* patient counts were approximated using the interval at form A05 recording (official withdrawal). *Other / Missing* represents those not having taken the main maternal interview form, (V#A)

|  | Visit 1 | Visit 2 | Visit 3 | Visit 4 (Delivery) | Post Delivery |
| --- | --- | --- | --- | --- | --- |
| **Present** (V#A) | 10,028 | 9,412 | 9,217 | 7,167 | 9,430 |
| **Withdrawn** | 7 | 167 | 213 | 276 | 62 |
| **Other or Missing** | 2 | 407 | 489 | 2,595 | 608 |

**Figure 3:**
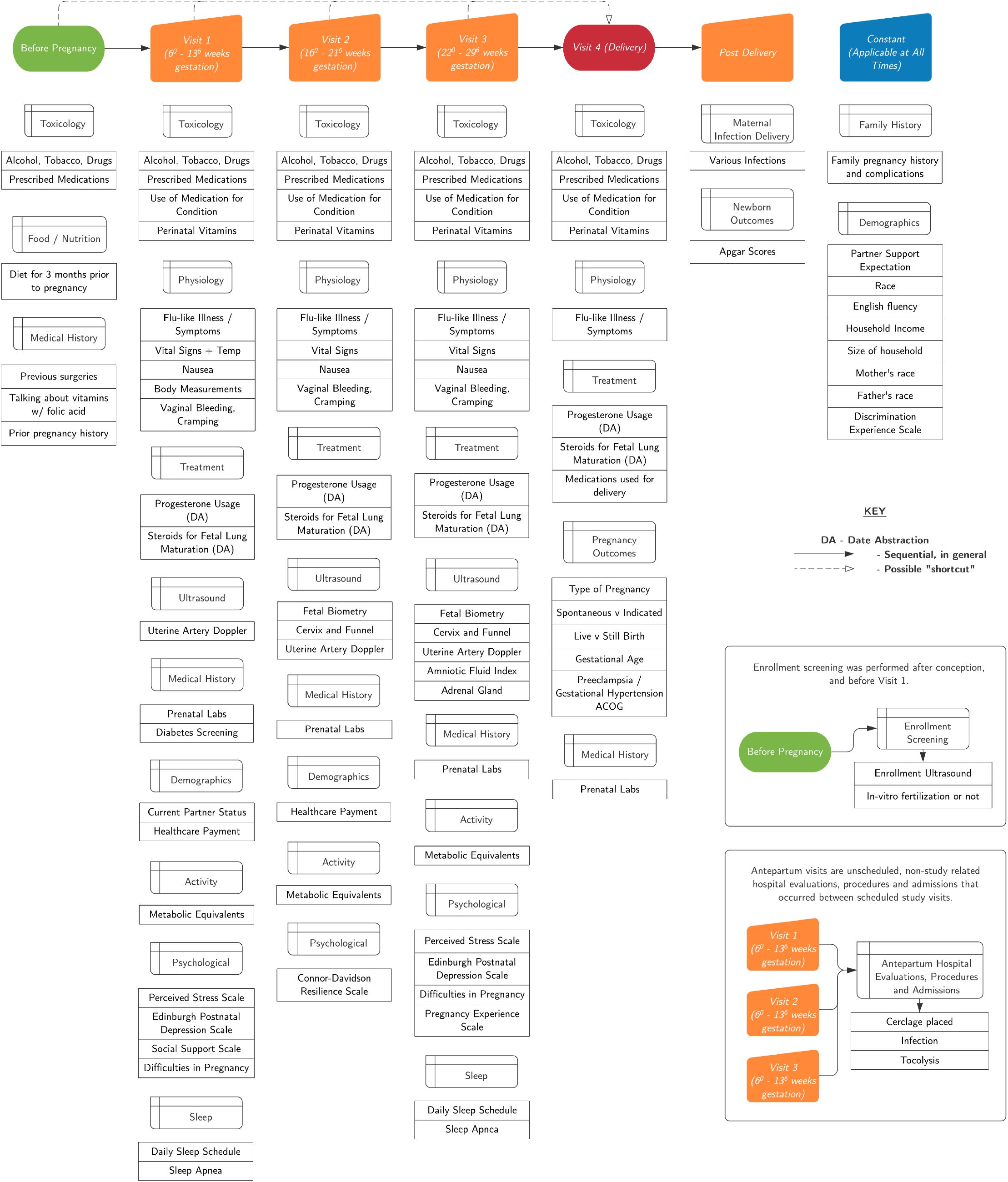
nuMoM2b Processed Data Timeline

## 6 Filtering Group Breakdown

The following is a breakdown of the filtering groups our team decided upon at the time of writing. These groups are subject to change in name and organization, and simply serve to understand and organize the data in a simpler and more manageable fashion. Each group below contains a description of itself, the total number of features marked belonging to said group, the number of features used in layer 1 and layer 2, the files which comprise the group, and a general description of the features that were dropped from out modeling. Following each description, shown is a table of the features both compiled and used as-is. Features used as-is are at the top of the table. Compiled features – those constructed from other variables in the data – follow, and are surrounded by horizontal lines and bolded. Those below a compiled variable are used to construct it, using the rule listed next to the compiled feature. The column **NAME** represents the original or compiled variable name. **FILE** represents the file in the data from which the original feature comes from. **RULE** represents which filtering rule was used to compile a group of features. **TEMPORAL** represents at which point in the timeline this feature is relevant. TEMPORAL ranges from -1 to 5, where 1 to 4 represent Visits 1 to 4 (delivery), -1 represents “applies at all time”, 0 represents before pregnancy, and 5 represents post-pregnancy. **IMPUTE** represents which rule was used for imputation. **MISSING** represents the missingness value for each original feature. **DESCRIPTION** is a shortened description from the original data set of each variable.

### 6.1 Family History

The family history filtering compiled all of the questions regarding diagnoses of family members related to diabetes, blood clotting disorders, pregnancy complications, heart disease, and hypertension. The 9 pregnancy-related conditions were grouped into 3 categories:

Spontaneous

- Early or preterm rupture of the membranes
- Spontaneous preterm delivery (less than 37 weeks)

Indicated

- Delivery of a child more than 3 weeks before the expected due date
- Preeclampsia, eclampsia, toxemia or pregnancy-induced hypertension

Fetal Conditions

- Delivery of a child weighing less than 5 lb 8 oz (or 2500 grams)
- Stillbirth
- Delivery of an infant with a birth defect
- Other pregnancy complication

Layer 1 includes binary features indicating any presence of family history in the 3 categories. Layer 2 compiled a more detailed score in the 3 categories which aggregates a score based on the genetic proximity to the stated family member, based on the following scale: 1 for mother, 0.75 for sister, 0.5 for half-sisters or cousins.

**Total # Features:** 113

**Layer 1 # Features:** 3

**Layer 2 # Features:** 3

**Relevant Files:** V2A

#### Dropped Features

We chose to discard family history related to diabetes, blood clotting disorders, heart disease, and hypertension, in order to focus on pregnancy-related conditions. Family history does NOT include the patient’s own medical history.

**Table 4:**
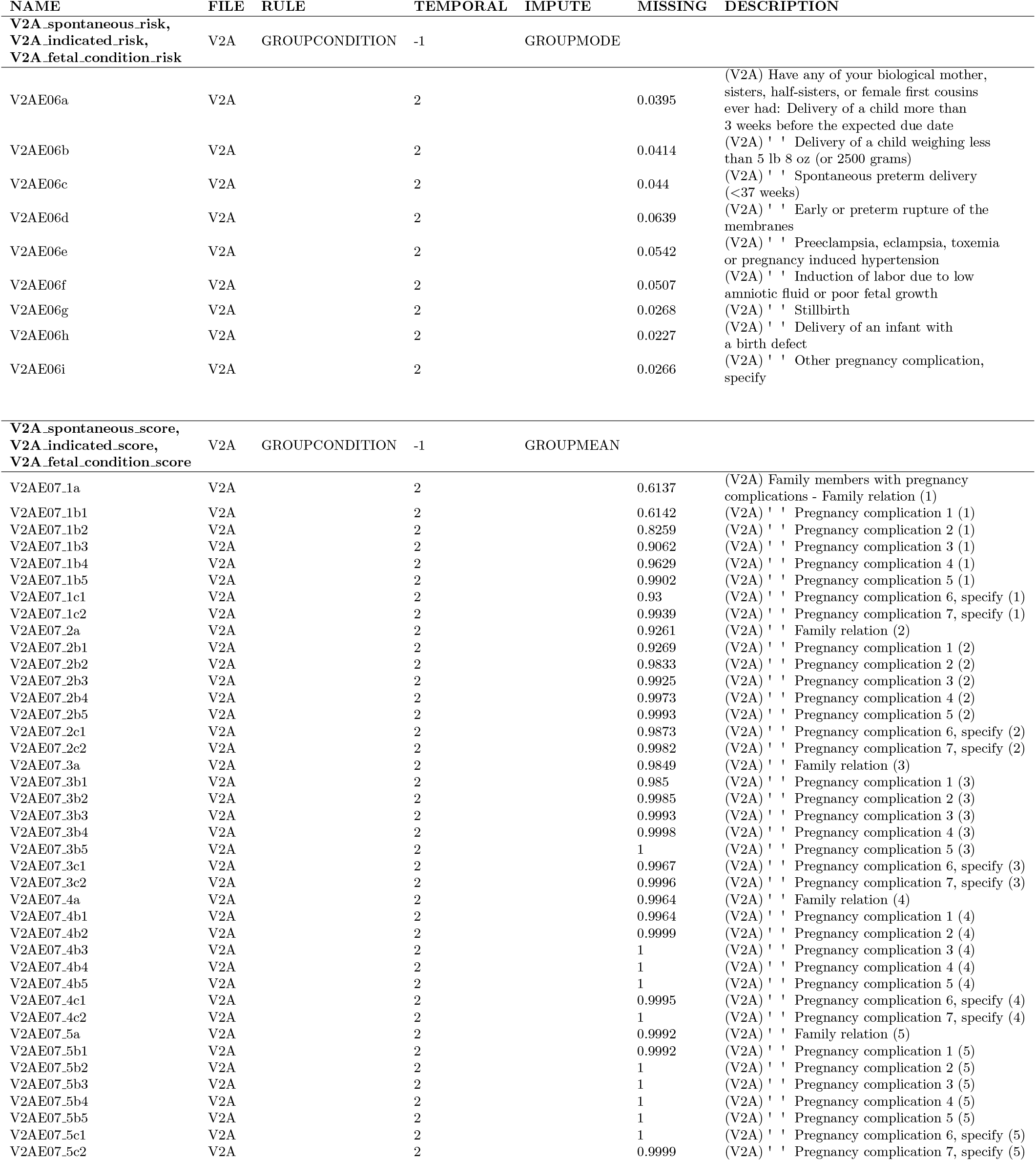
Family History Filtering

### 6.2 Toxicology

The toxicology filtering relates to questions and chart abstractions regarding any kind of drug use, prescription or non-prescription, by the patient. This includes questions regarding alcohol, tobacco usage, second-hand smoke, smoking cessation, illegal drugs, all prescription medications noted, vitamins, vaccines, and whether medication was being taken for a particular condition. The information used comes from both patient interviews and ancillary files, which corrected the medication information through consensus and chart review.

**Total # Features:** 806

**Layer 1 # Features:** 63

**Layer 2 # Features:** 63

**Relevant Files:** drugs in pregnancy (ancillary), VXX, V1A, V2A, V3A, V4A

#### Dropped Features

General questions on if a class of drug (non-prescribed) was used were kept in L1. Some detail was saved for L2, such as illegal drug use breakdown. Prescription drugs were organized back into drug categories originally used by the nuMoM2b team in order to reduce dimensionality (see form VXX Section C), perinatal vitamins were separated from the vitamins group due to feature compiled feature *Vitamin Multi Perinatal Folate*, covering that information as well. Metadata features were used to organize and preprocess data, and then dropped from modeling. “Medication taken for condition” features in VXX are dropped as prescribed drugs (as mentioned earlier) and “condition noted” features from VXX in medical history filtering cover said information.

**Table 5:**
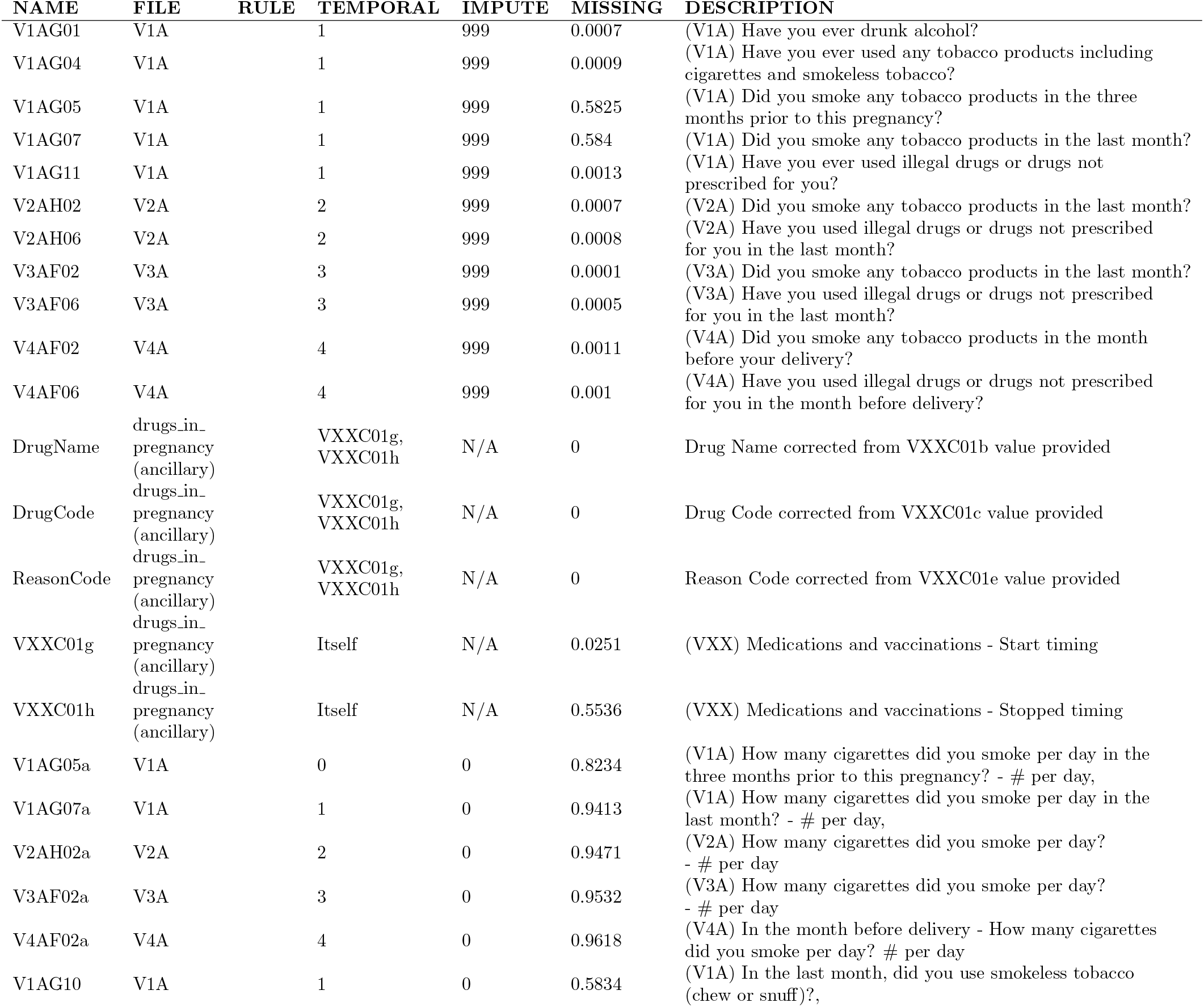
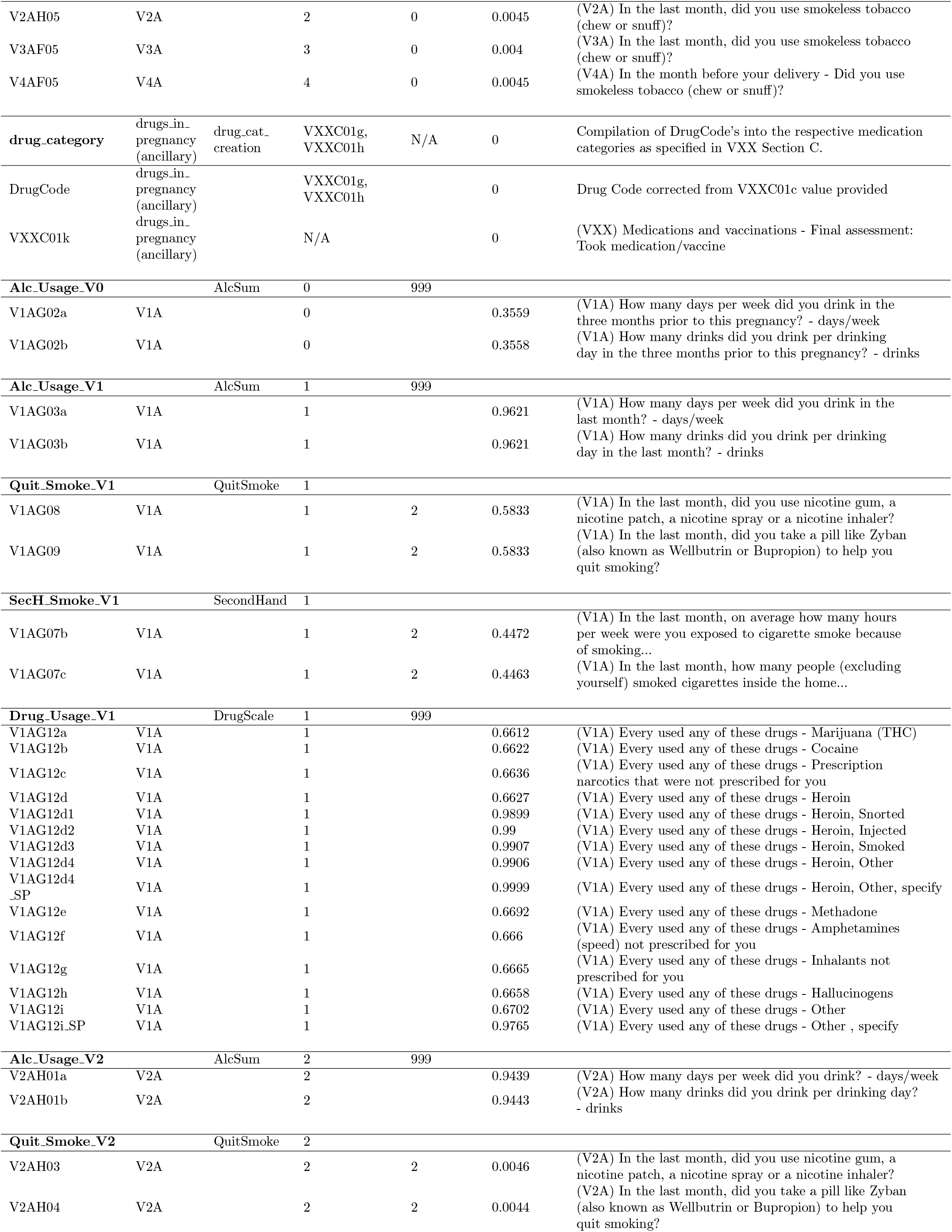
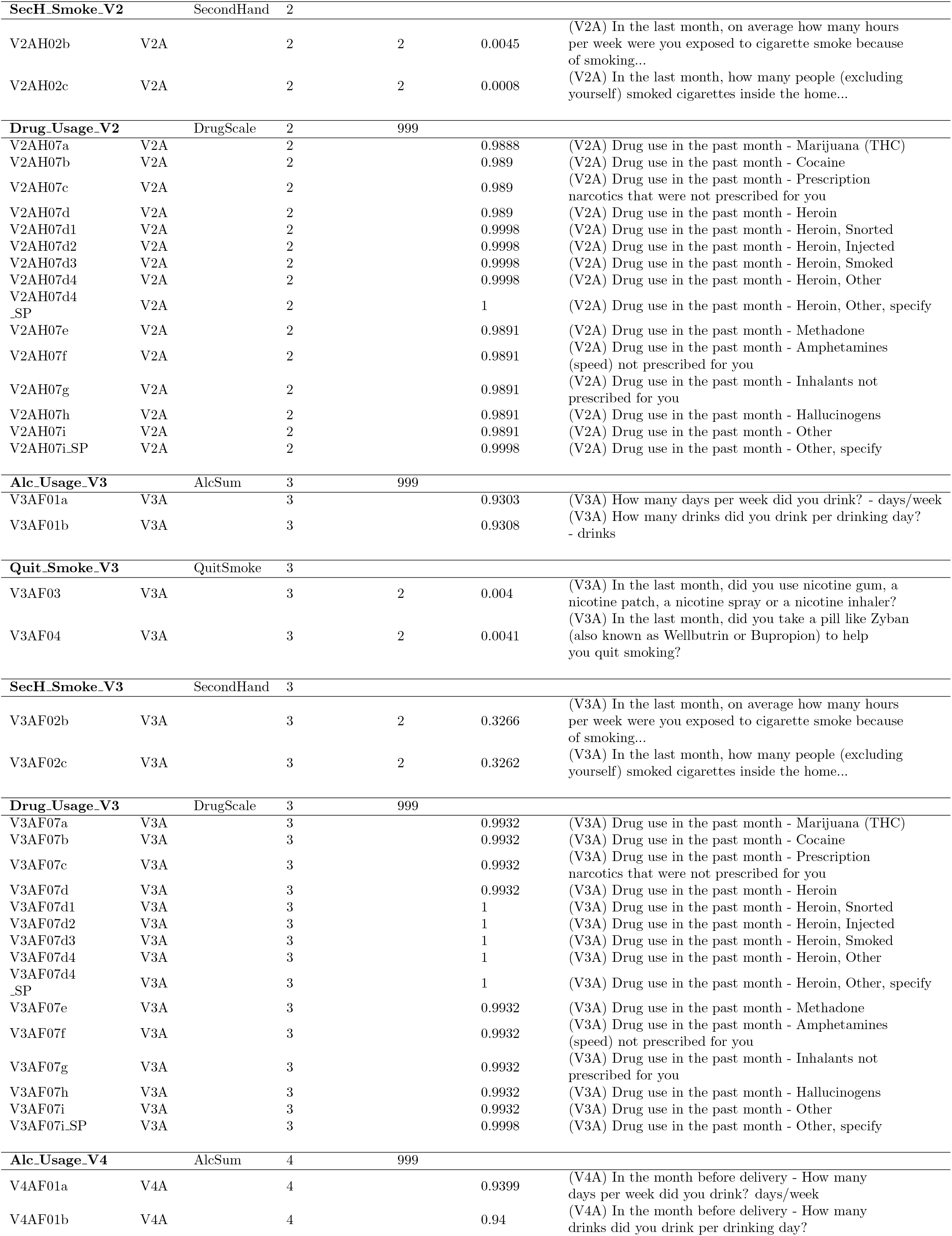
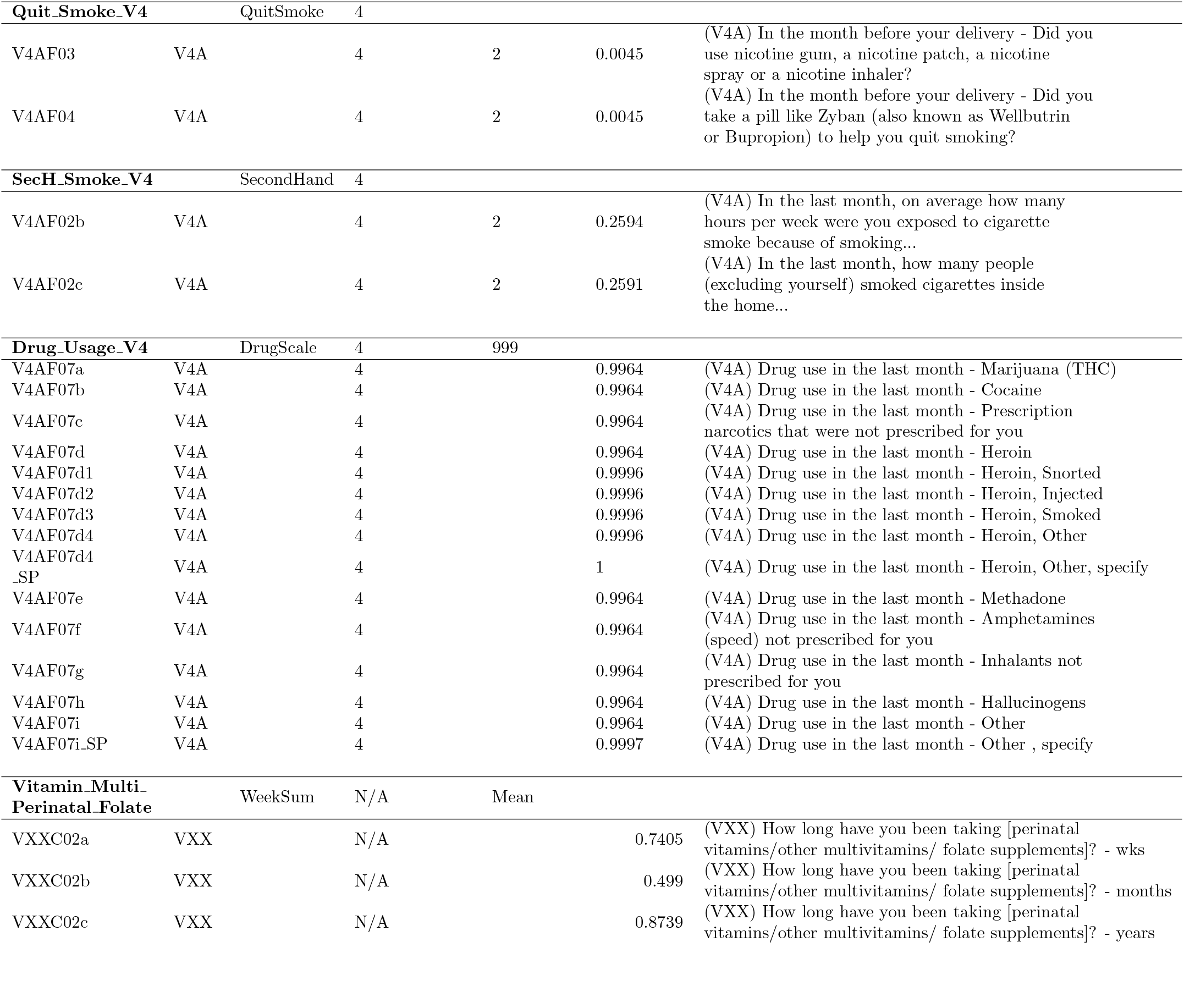
Toxicology Filtering

### 6.3 Psychological

The psychological filtering compiled all of the features relating to psychological health and wellbeing, including previous treatment for mental health conditions in addition to surveys about factors related to depression, stress, resiliency and social support at the 3 visits.

**Total # Features:** 172

**Layer 1 # Features:** 8

**Layer 2 # Features:** 8

**Relevant Files:** CMA, V1A, V2A, V1C, V1E, V1G, V1H, V2I, V3A, V3C, V3E, V3J

#### Dropped Features

We chose to discard features related to prenatal and postpartum treatment for mental health conditions, in the case that some conditions were not diagnosed, to focus on the current state of mind throughout the pregnancy as measured by the surveys.

**Table 6:**
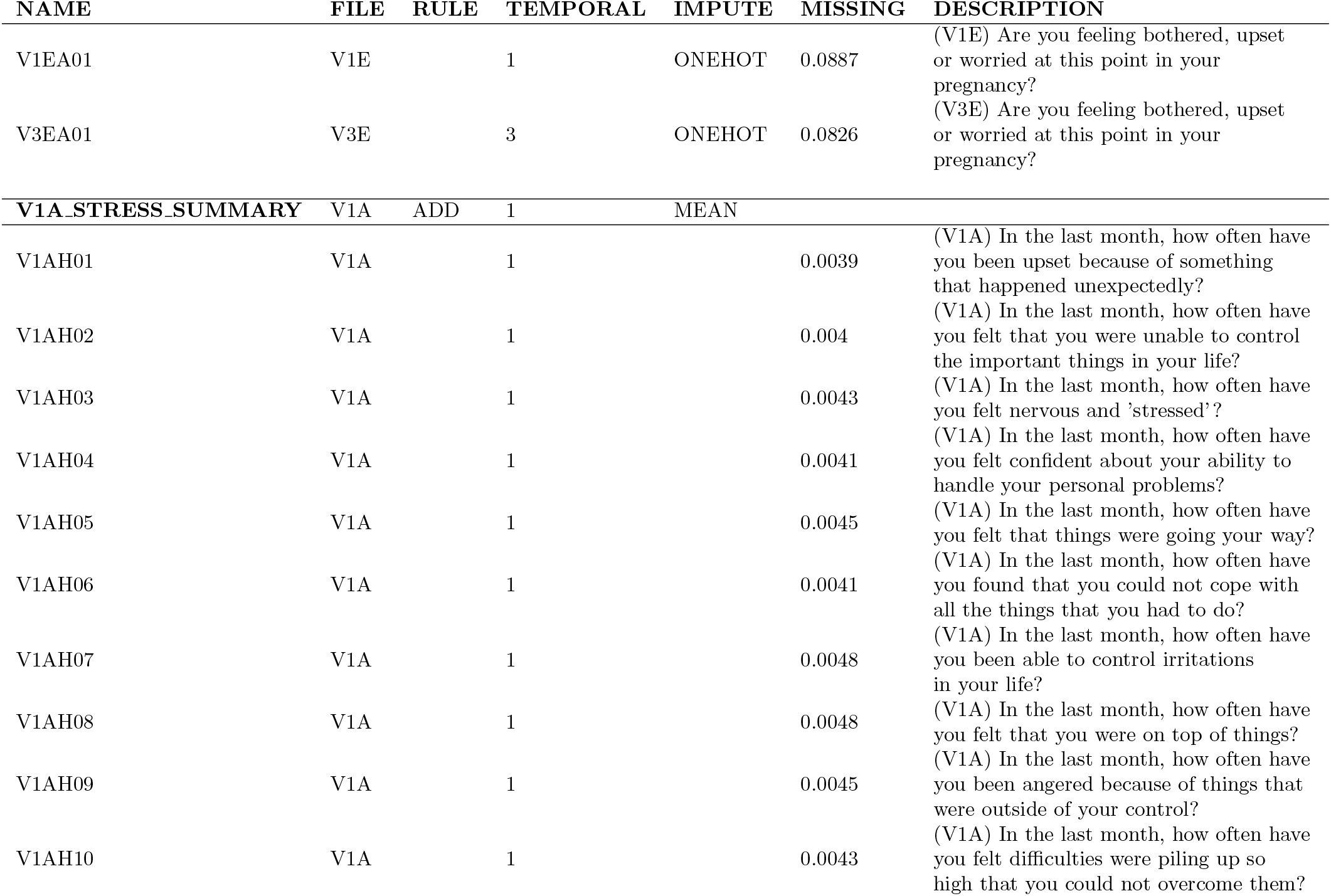
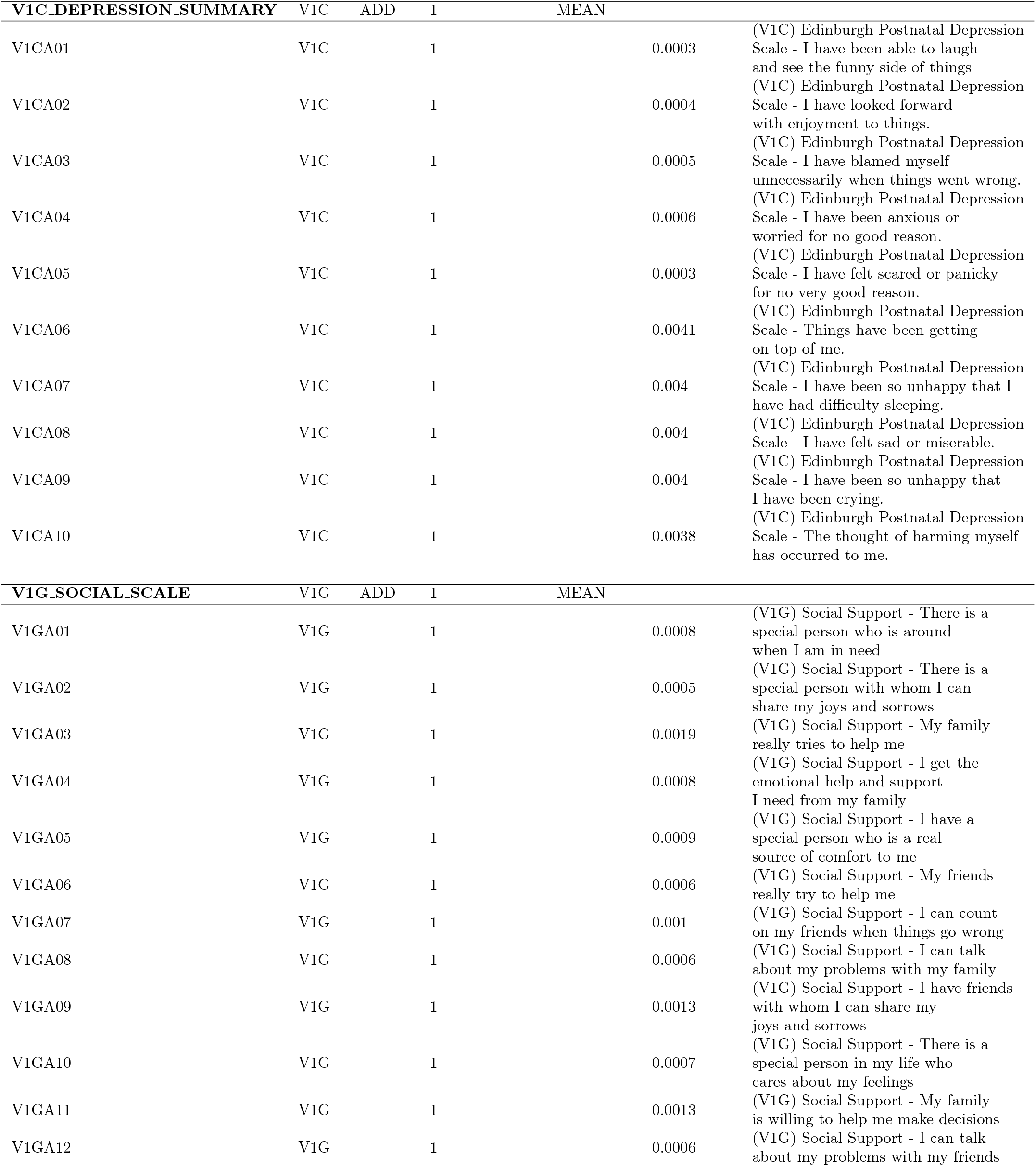
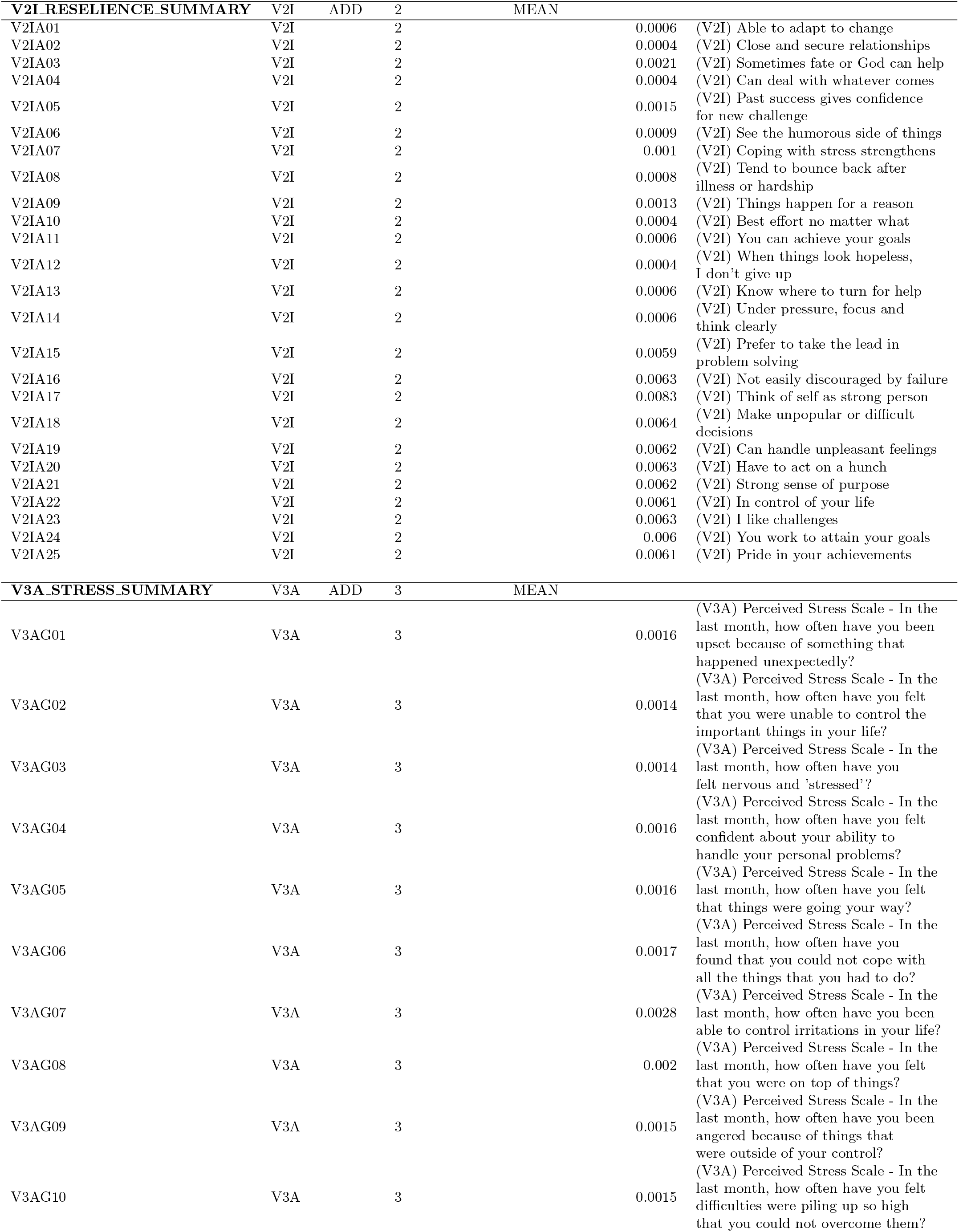
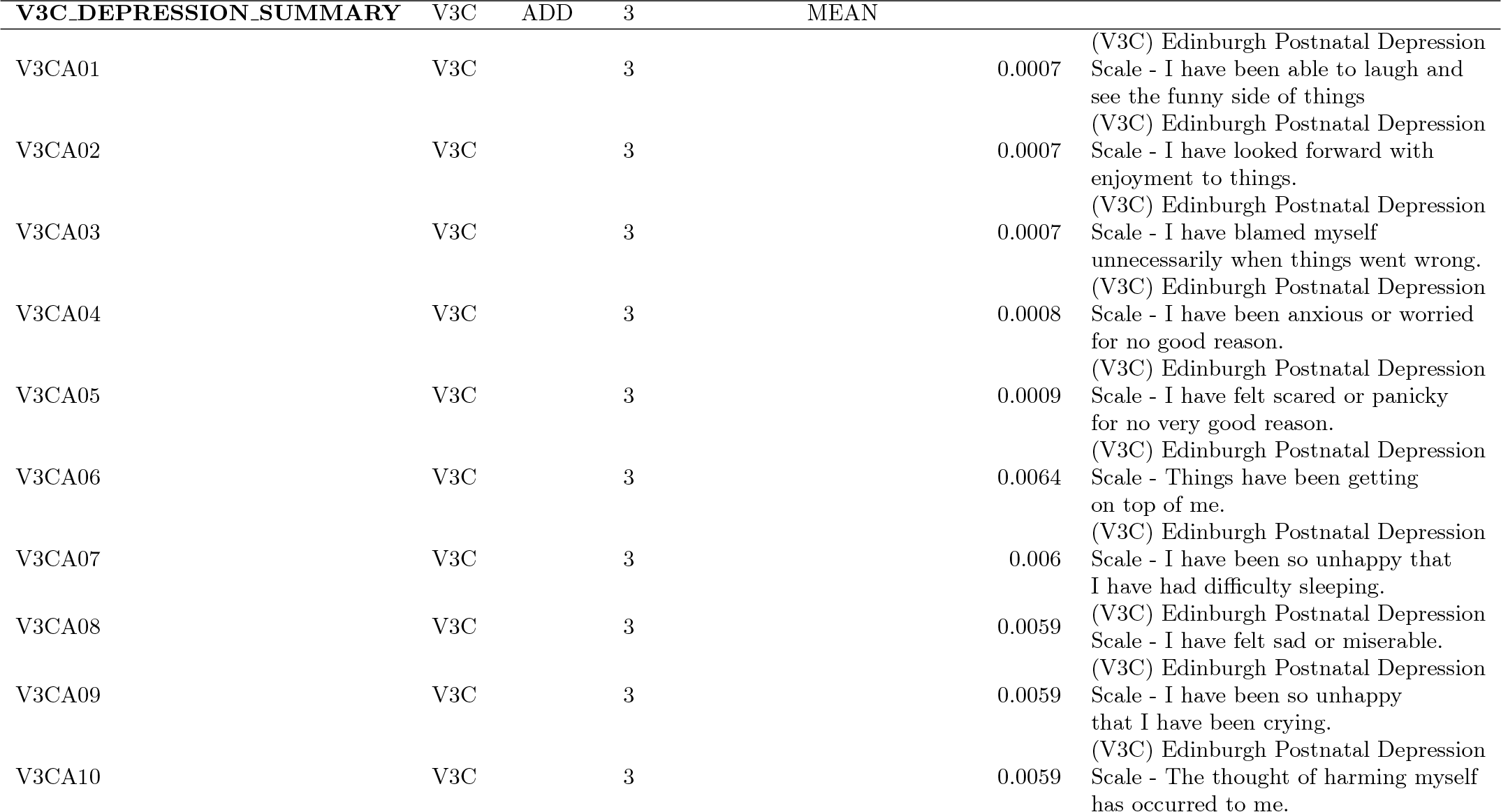
Psychological Filtering

### 6.4 Activity

The activity filtering related to details about participant physical activity prior to and during pregnancy. The activities were measured in METs, or metabolic equivalents, for Layer 1 and expanded upon in further detail, namely activity type, minutes, miles and duration in Layer 2. Activity restriction information available at V3 and V4 was also included, to account for influence on the MET score.

**Total # Features:** 131

**Layer 1 # Features:** 3

**Layer 2 # Features:** 22

**Relevant Files:** physical activity (ancillary), V1A, V2A, V3A, V4A

#### Dropped Features

We chose to discard features relating to any personal or care provider weight change goals, as these were suggestions that would merely be reflected in the actual activity data.

**Table 7:**
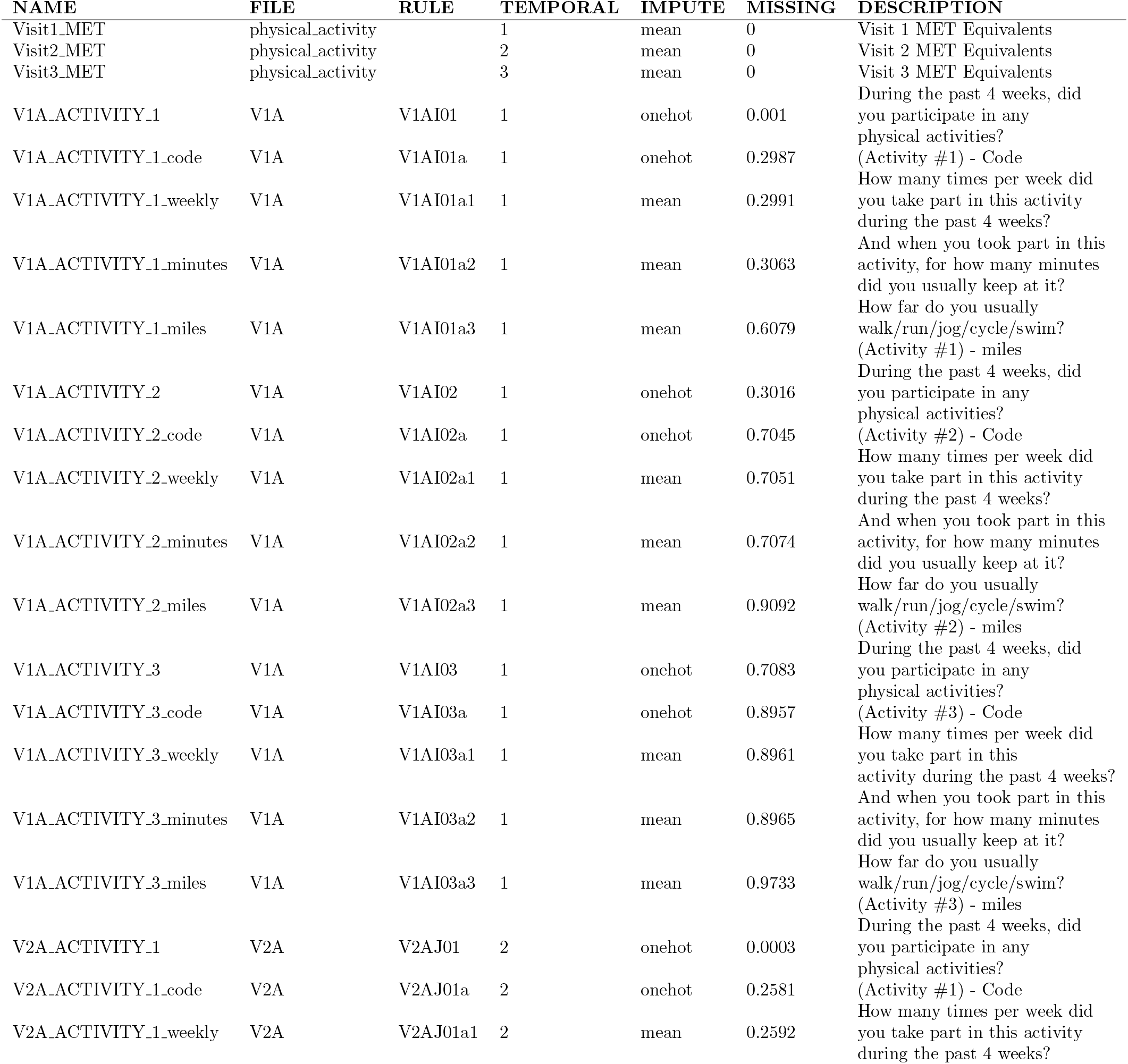
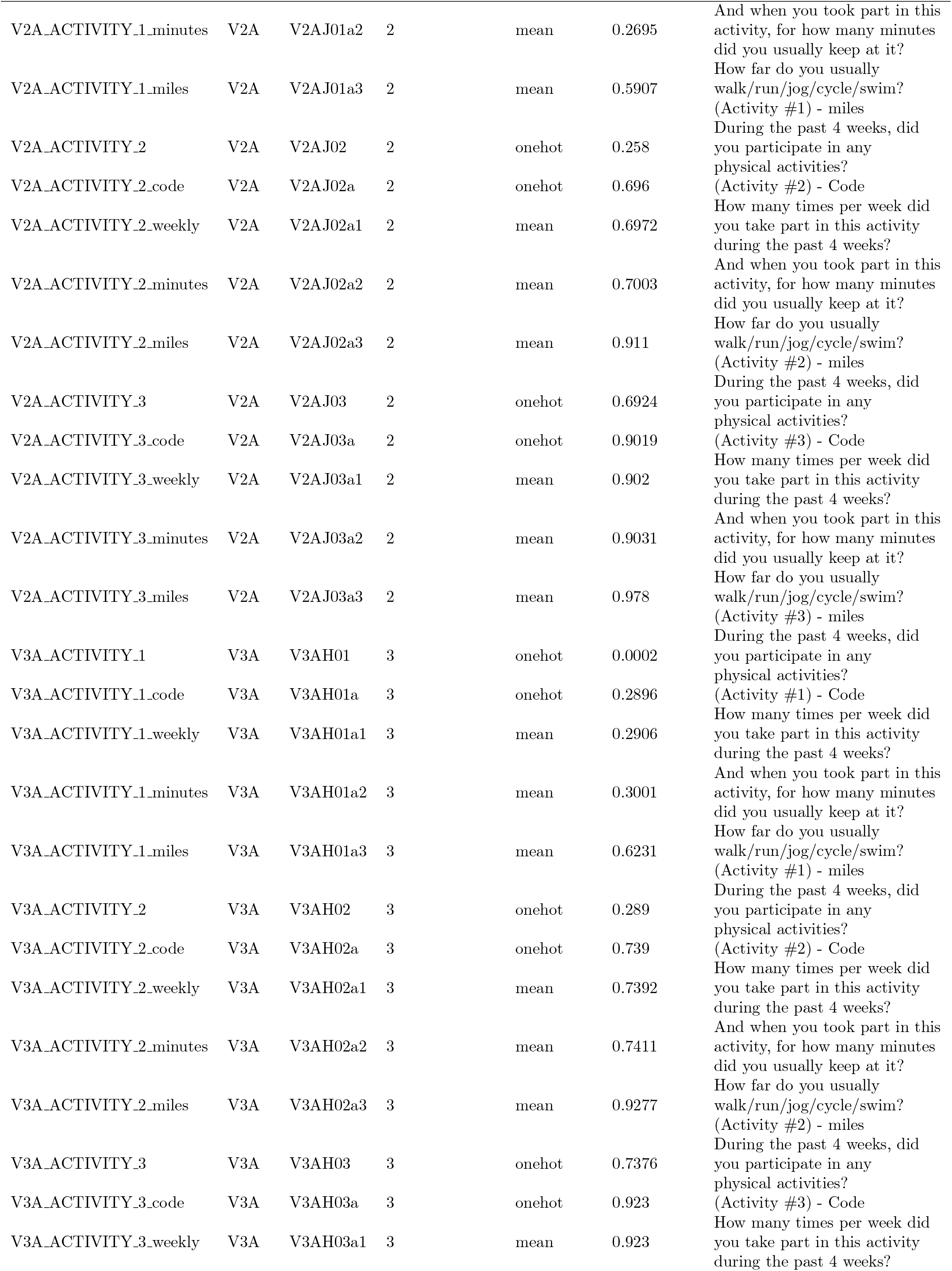
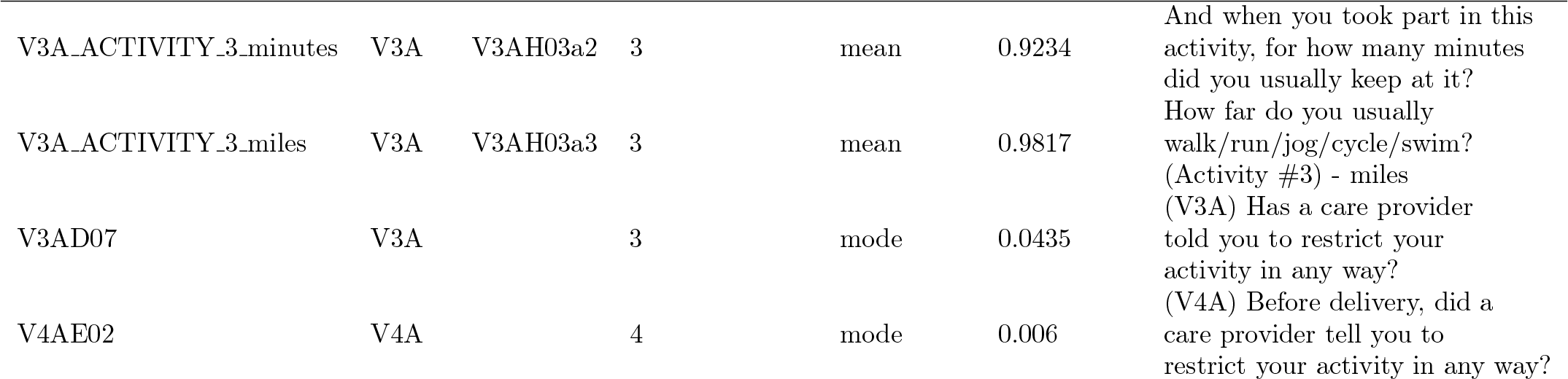
Activity Filtering

### 6.5 Demographics

The demographics filtering includes details about social factors including education and marital status, in addition to the more medically relevant factors of race, healthcare access and stress levels related to partner support.

**Total # Features:** 170

**Layer 1 # Features:** 21

**Layer 2 # Features:** 22

**Relevant Files:** V1A, V2A

#### Dropped Features

We chose to discard redundant features such as the racial background of the mother’s parents since that information is inherently stored in the mother’s race herself. Social constructs like ethnicity and perceived race (by others) were not deemed medically relevant, so those were also removed. Experiences in discrimination would further account for some of their social effects. We also believed that expectations for partner support was a more relevant proxy of social support than questions about the nature of the relationship such as length of relationship.

**Table 8:**
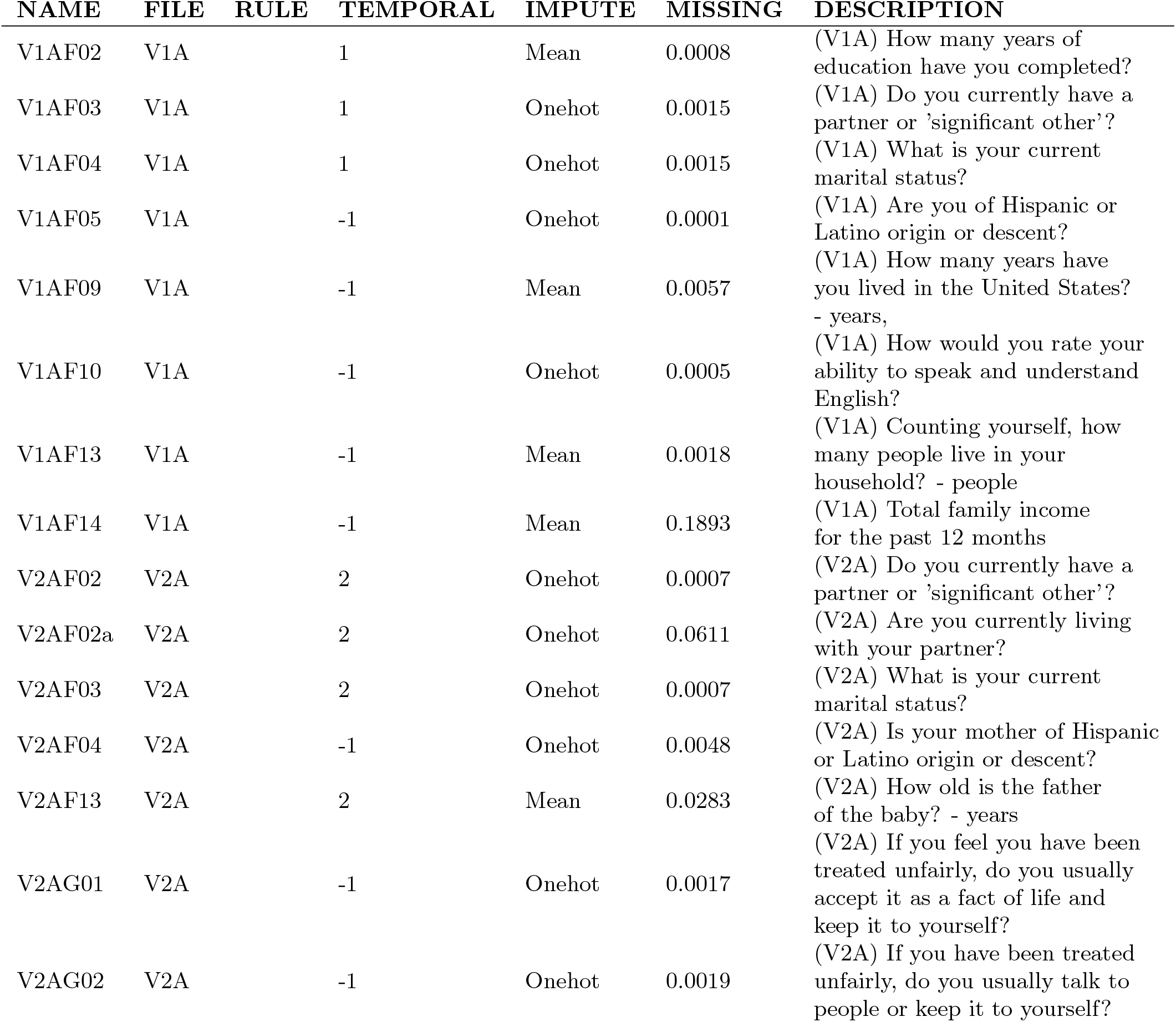
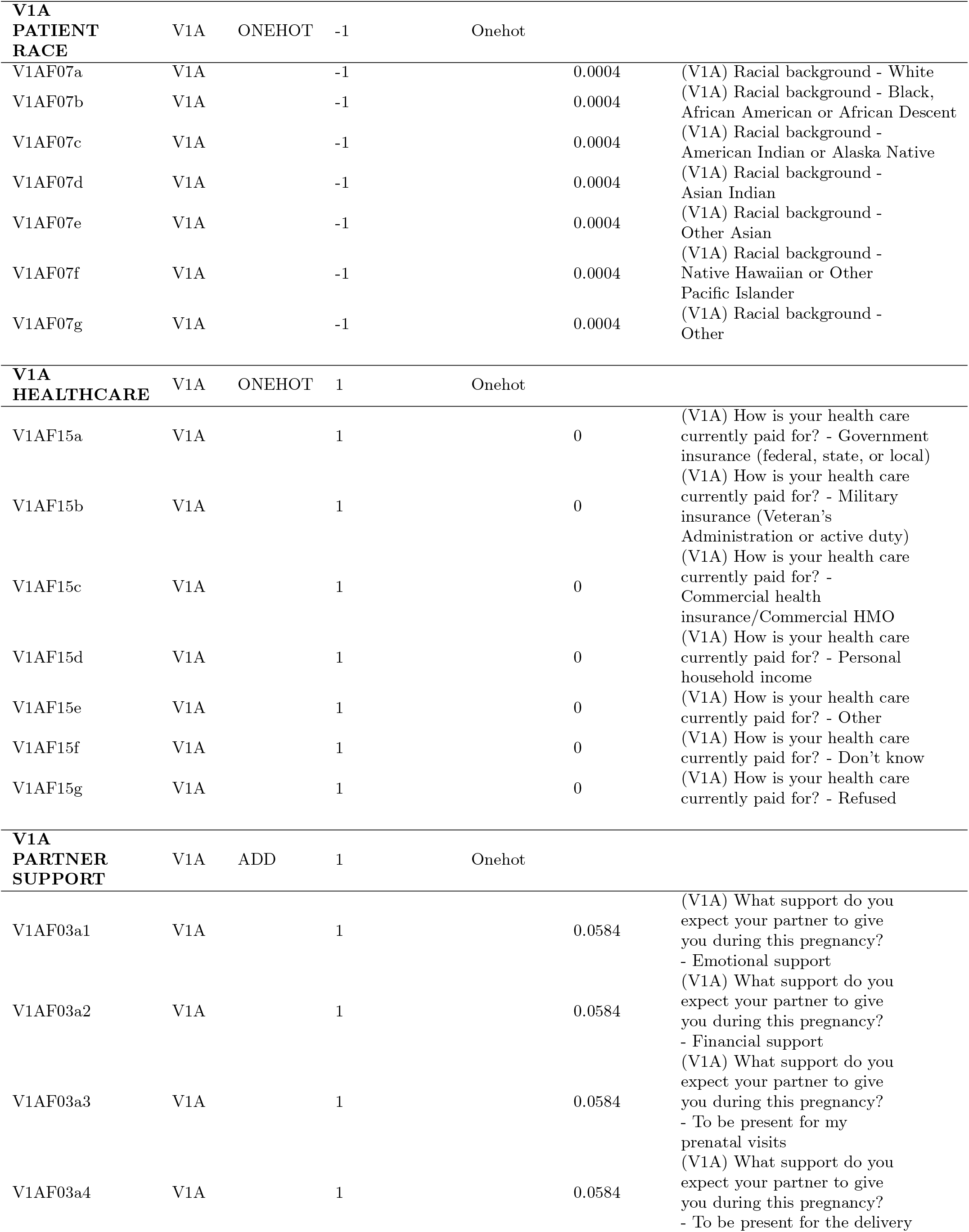
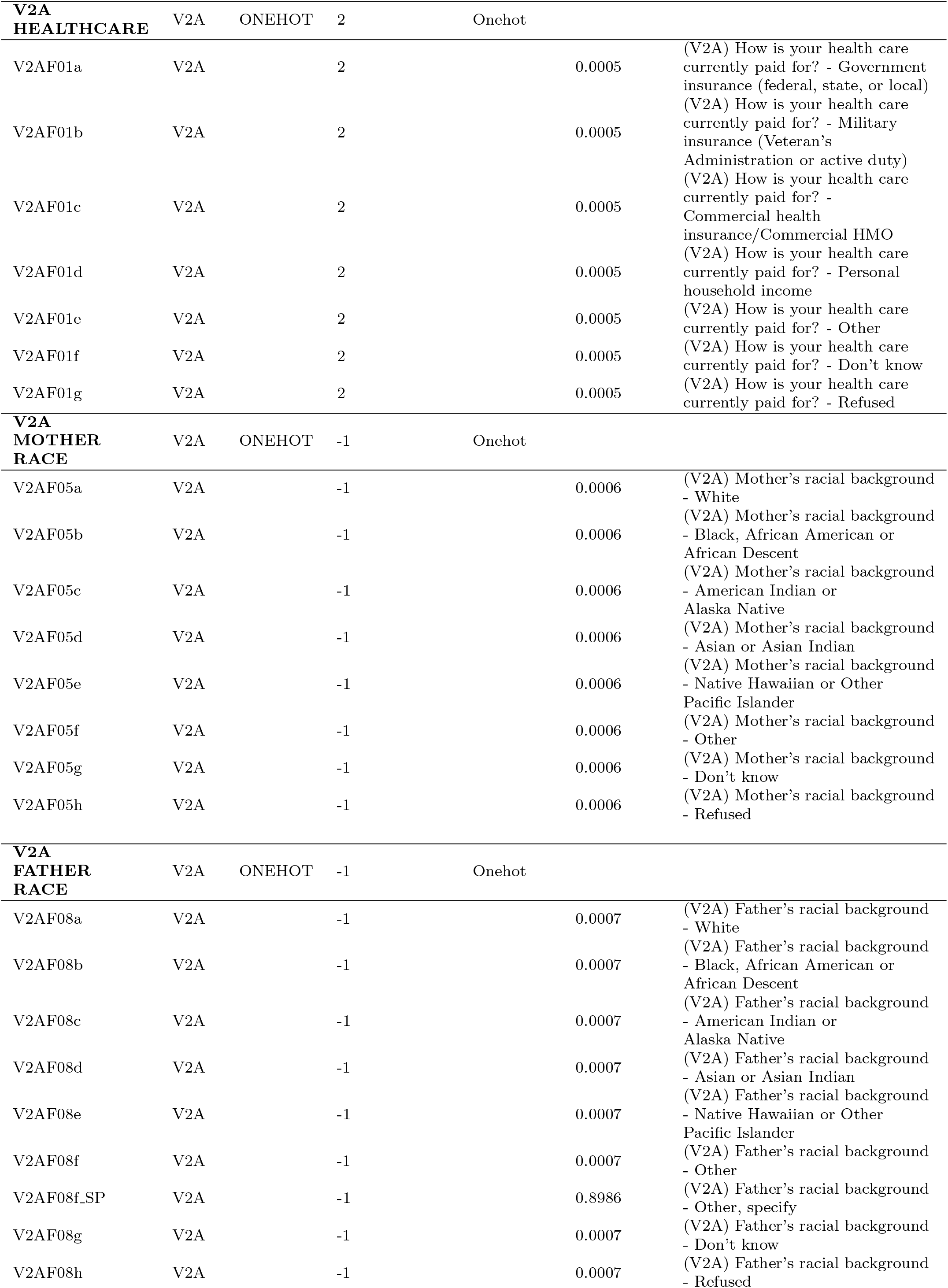
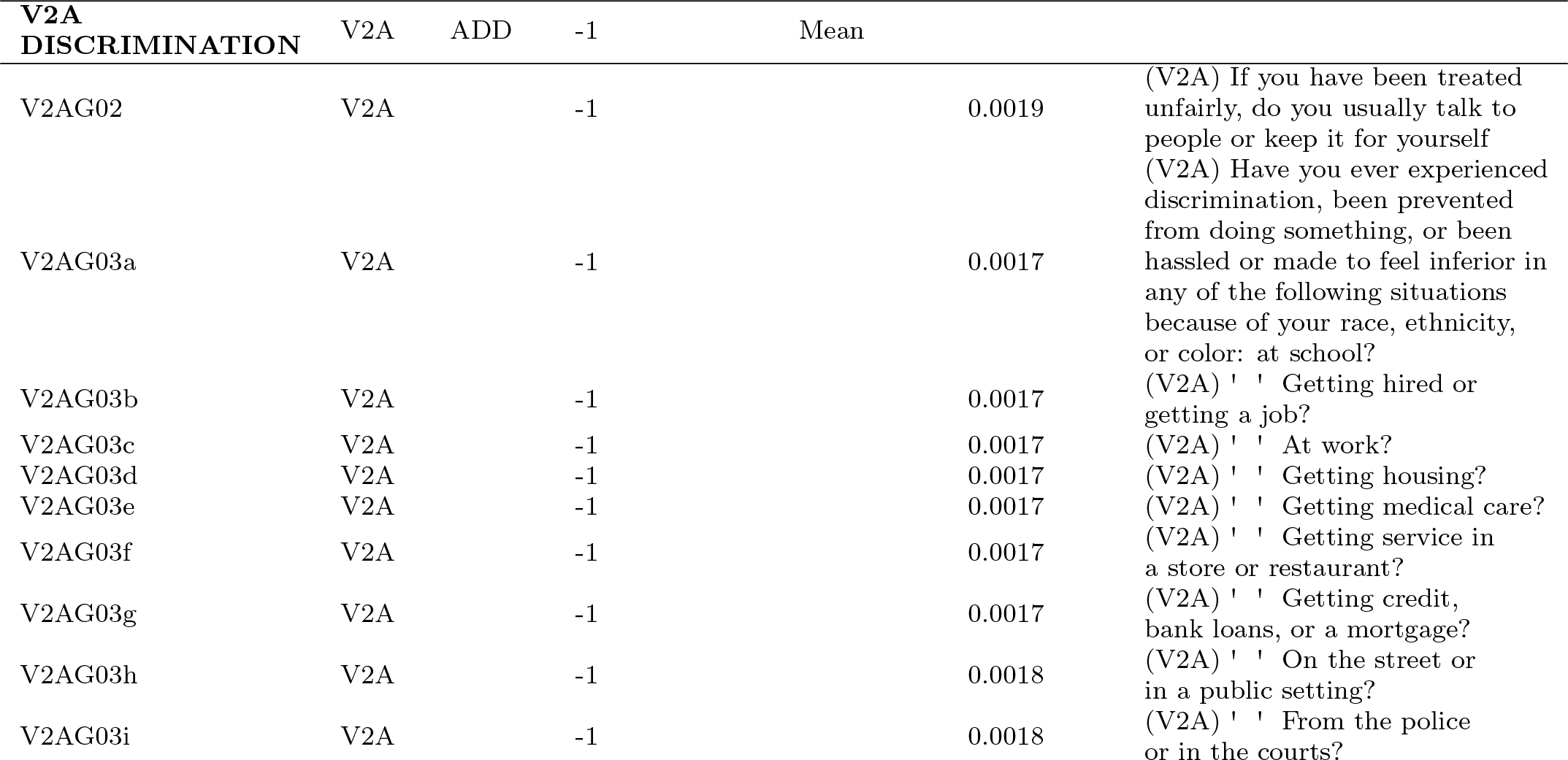
Demographics Filtering

### 6.6 Physiology

The physiology filtering includes questions regarding the current and recent physiological health and questioning of the patient. This includes factors such as temperature, weight, body measurements, flu-like symptoms, nausea, blood pressure, and recent evidence of contractions or vaginal bleeding.

**Total # Features:** 115

**Layer 1 # Features:** 38

**Layer 2 # Features:** 38

**Relevant Files:** CMA, CMB, Demographics (ancillary), V1A, V1B, V2A, V2B, V3A, V3B, V4A

#### Dropped Features

For this group, we chose to exclude features that were detail oriented to questions that were asked earlier in the questionnaire. For example, details on flu-like illness symptoms were dropped, while the general question on if any such illness was present was kept, as the general question implicitly covers the presence of a response in the details that follow. Certain details were added back within L2, such as contraction and vaginal bleeding times. Multiple measurements of the same region were averaged into a single feature. Maximum, rather than average, blood pressure and temperature on admission were kept as they were deemed a better representation of potential risk or medical issues. Metadata features were used to organize data and later dropped.

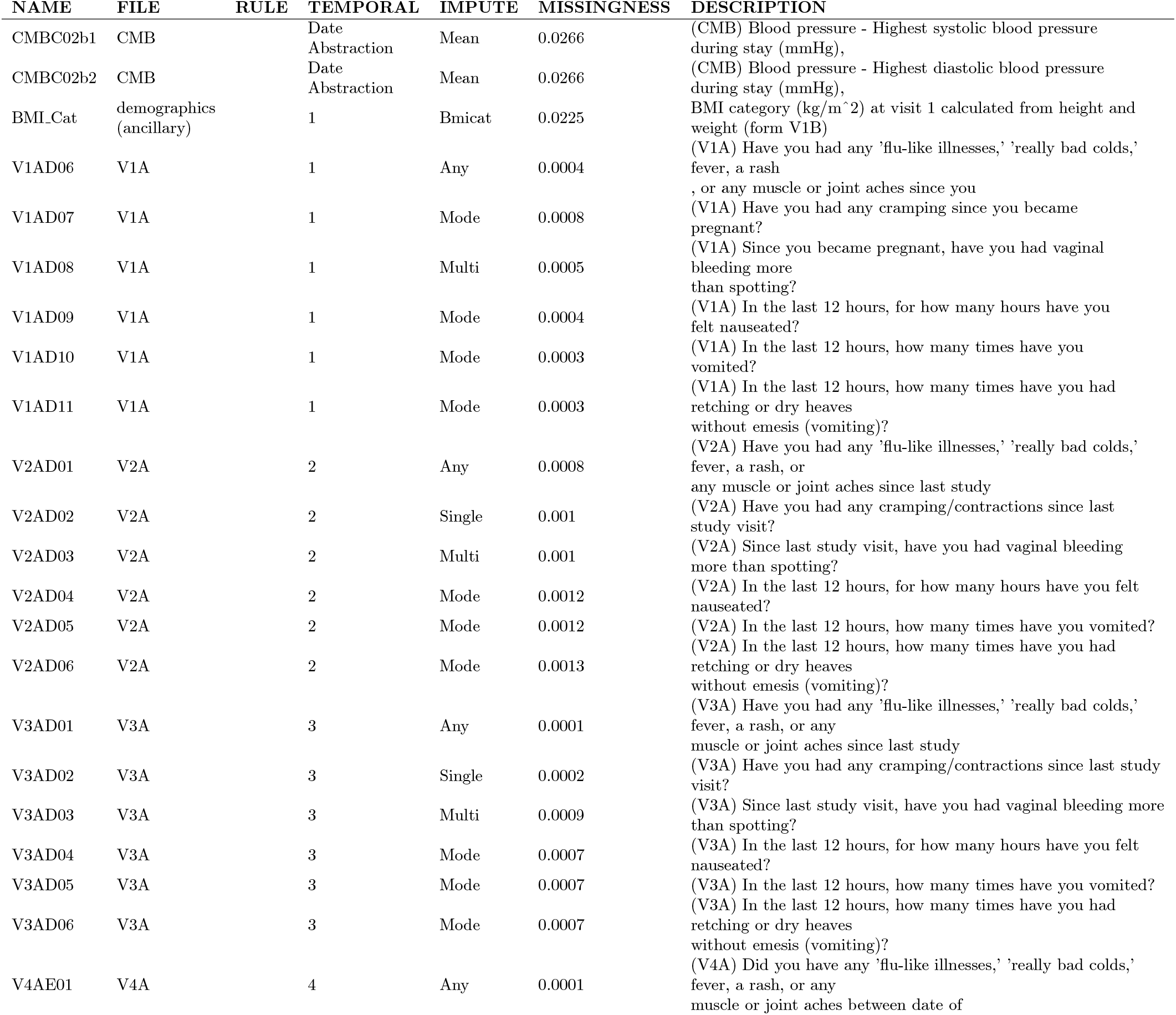
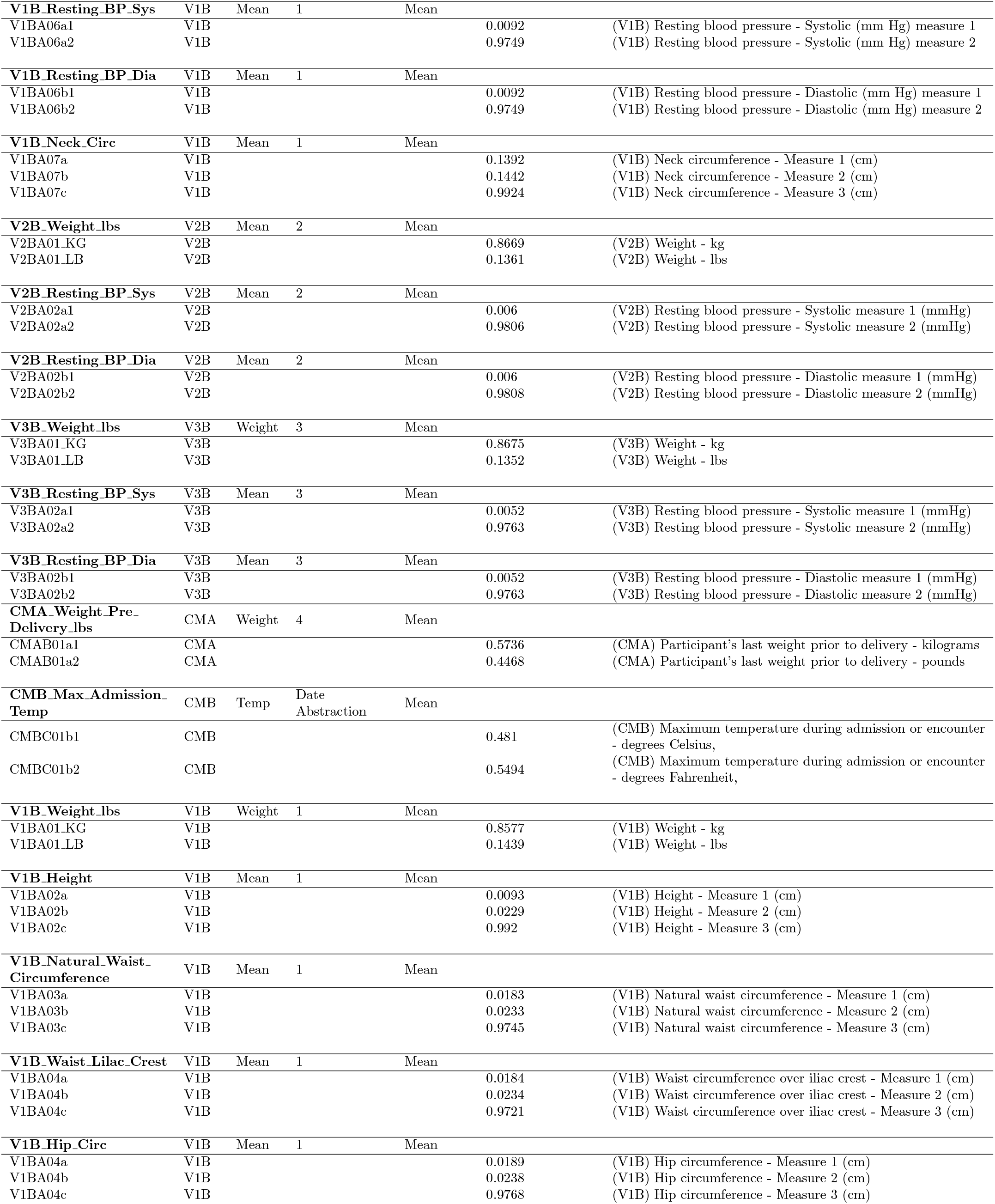

### 6.7 Ultrasound

The ultrasound filtering group contains all features located within the ultrasound related files and chart abstractions. This includes fetal measurements, uterine artery analysis, adrenal gland study, noted conditions, amniotic fluid information, and more.

**Total # Features:** 542

**Layer 1 # Features:** 59

**Layer 2 # Features:** 131

**Relevant Files:** CUA, CUB, S02, U02, U1C, U2A, U2B, U2C, U3A, U3B, U3C, U3D

#### Dropped Features

For this group, there were many excess features that were dropped (100% missingness), for noted abnormalities. Abnormality code was the only feature used to represent fetal abnormalities. There was abundant metadata relevant only for either organizing data or populating other features and therefore redundant, which was dropped. Details for conditions were left for Layer 2, as were detailed measurements in the uterine artery.

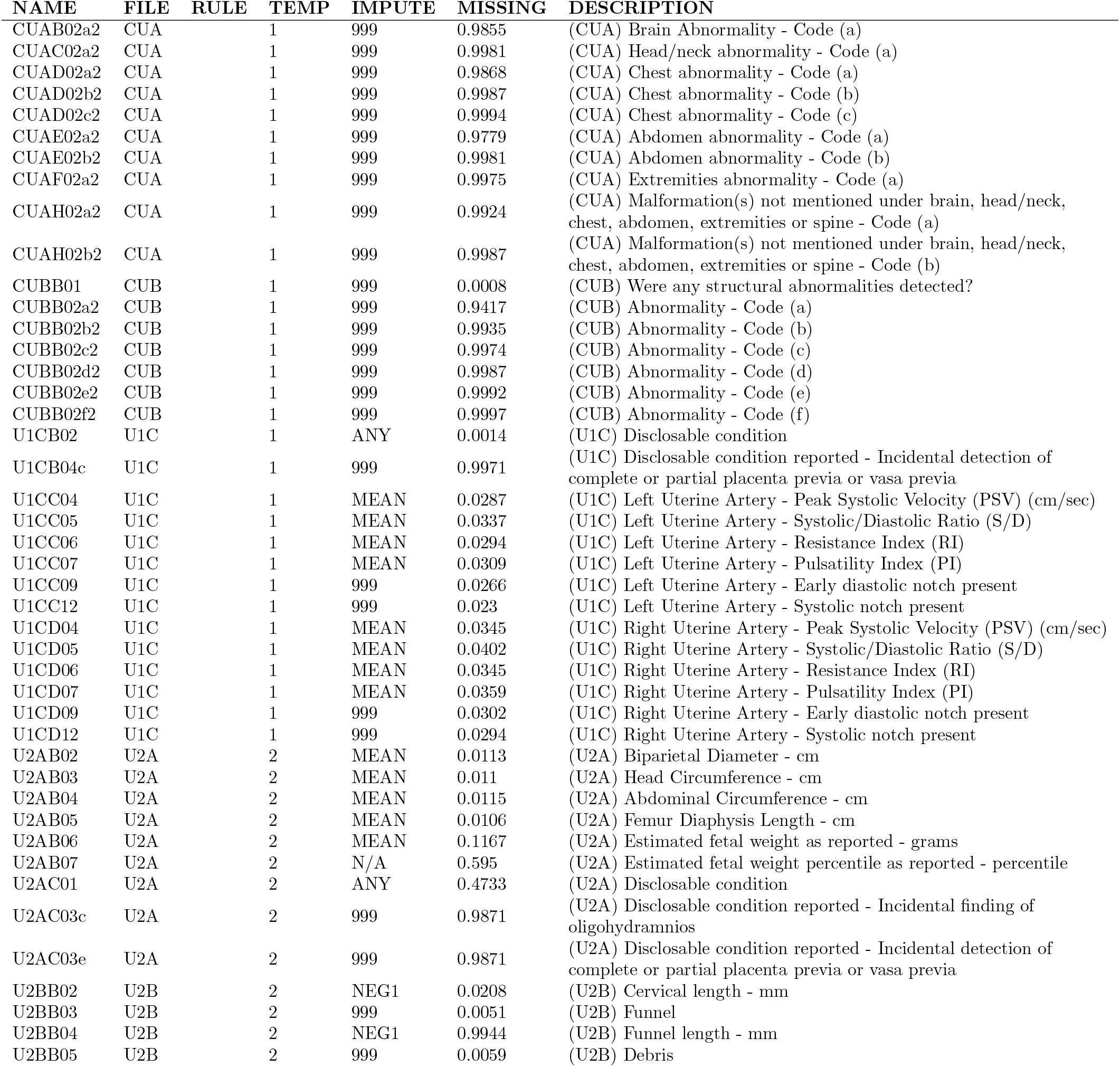
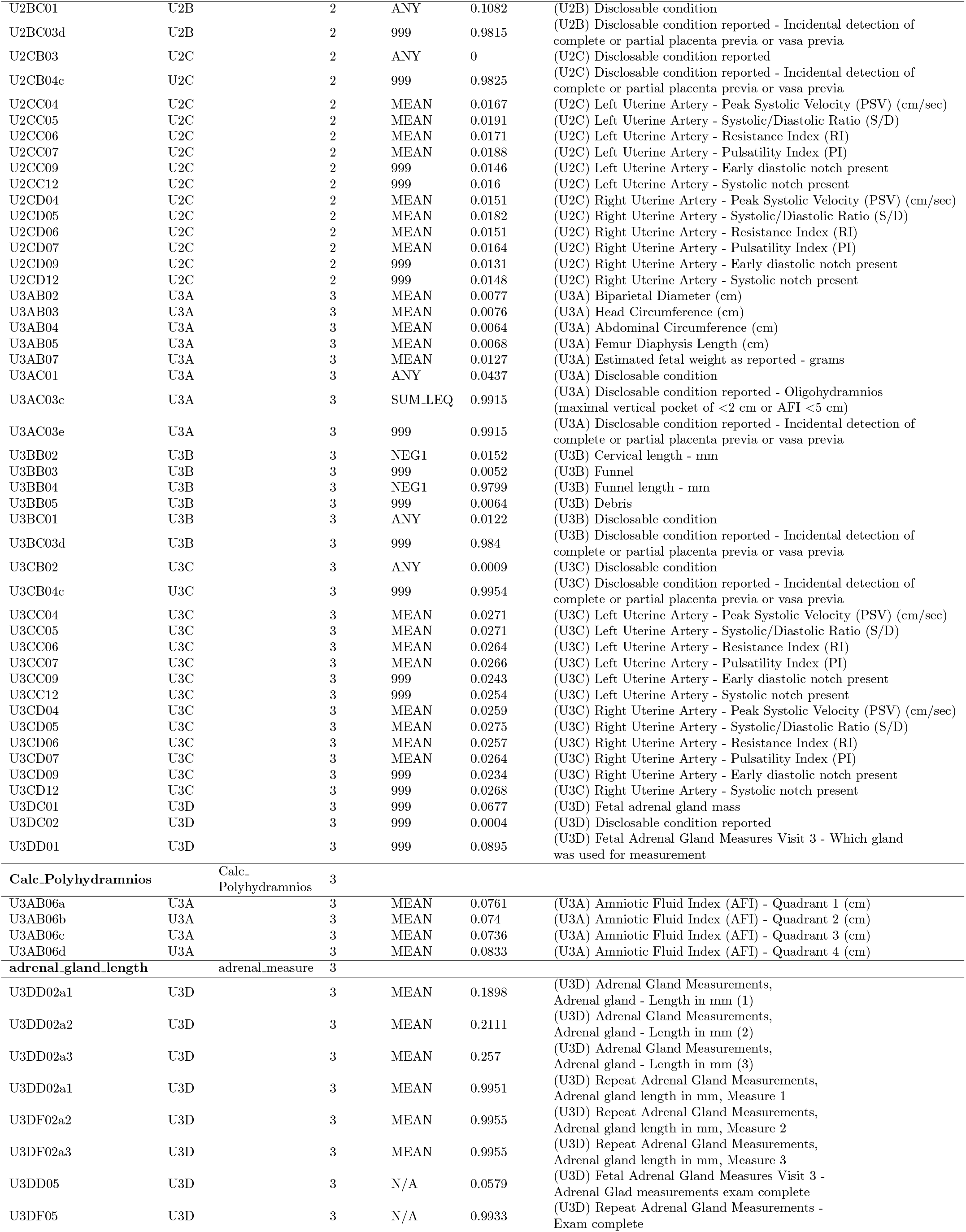
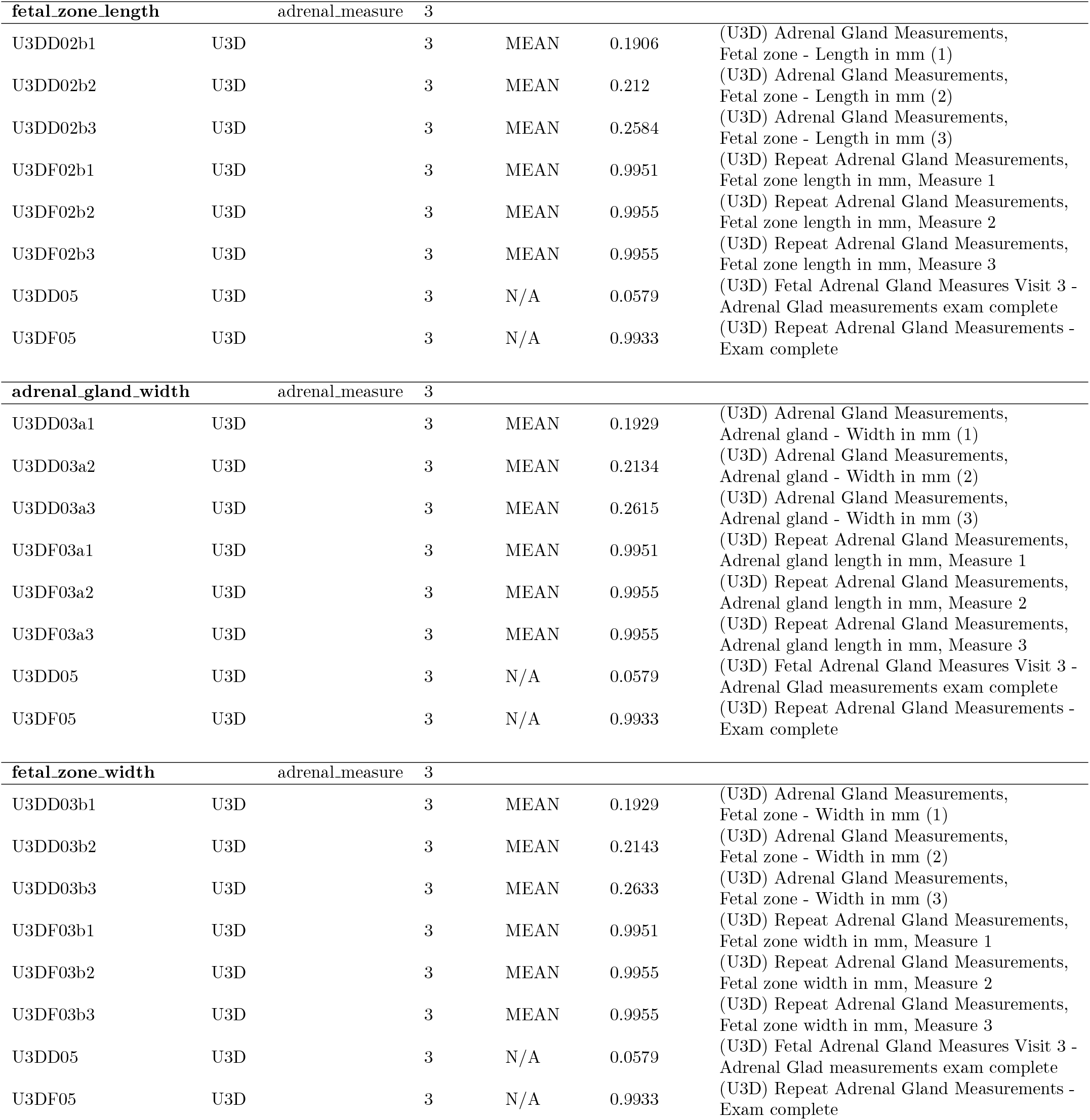
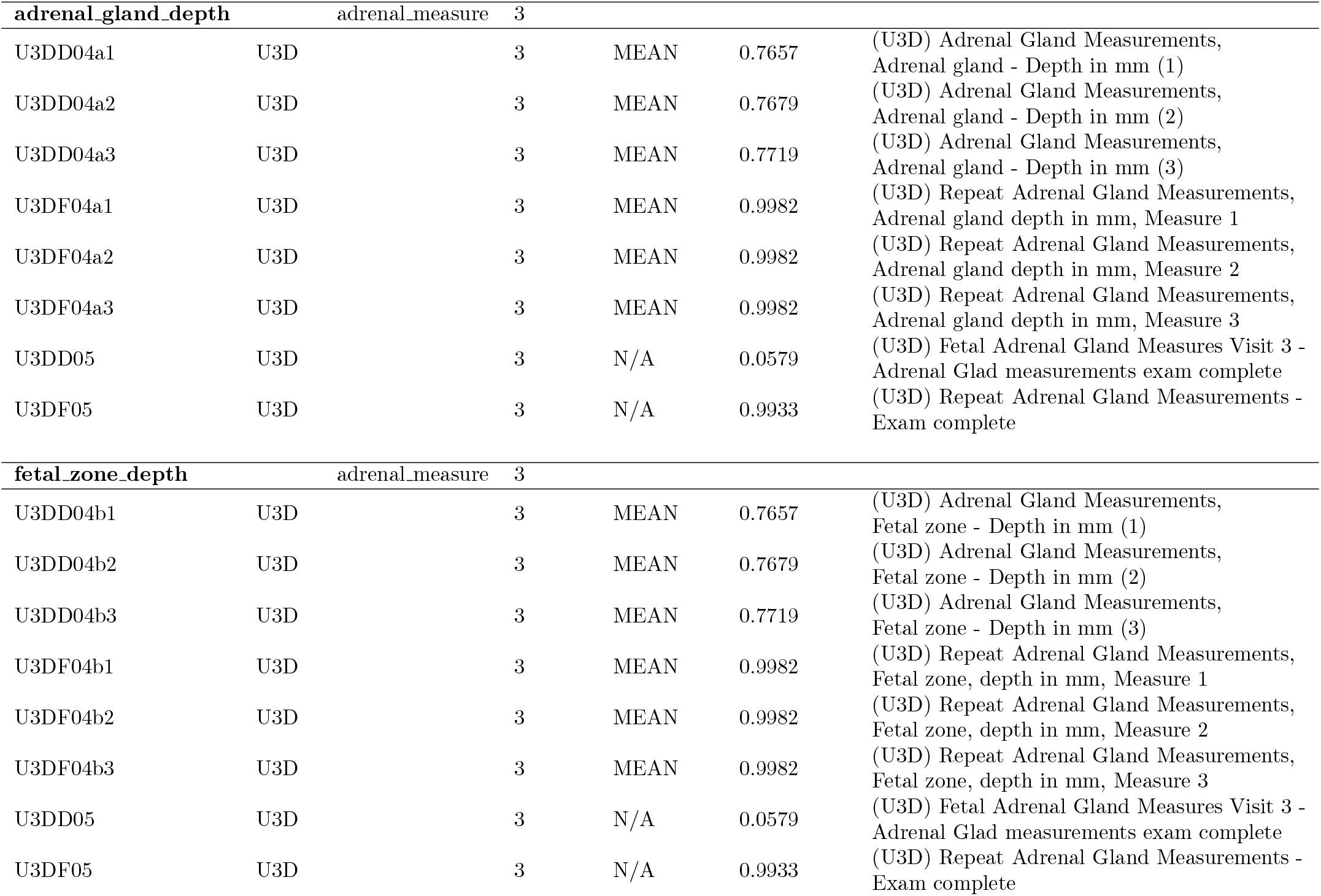

| NAME | FILE | RULE | TEMP | IMPUTE | MISSING | DESCRIPTION |
| --- | --- | --- | --- | --- | --- | --- |
| CUAB02a2 | CUA |  | 1 | 999 | 0.9855 | (CUA) Brain Abnormality - Code (a) |
| CUAC02a2 | CUA |  | 1 | 999 | 0.9981 | (CUA) Head/neck abnormality - Code (a) |
| CUAD02a2 | CUA |  | 1 | 999 | 0.9868 | (CUA) Chest abnormality - Code (a) |
| CUAD02b2 | CUA |  | 1 | 999 | 0.9987 | (CUA) Chest abnormality - Code (b) |
| CUAD02c2 | CUA |  | 1 | 999 | 0.9994 | (CUA) Chest abnormality - Code (c) |
| CUAE02a2 | CUA |  | 1 | 999 | 0.9779 | (CUA) Abdomen abnormality - Code (a) |
| CUAE02b2 | CUA |  | 1 | 999 | 0.9981 | (CUA) Abdomen abnormality - Code (b) |
| CUAF02a2 | CUA |  | 1 | 999 | 0.9975 | (CUA) Extremities abnormality - Code (a) |
| CUAH02a2 | CUA |  | 1 | 999 | 0.9924 | (CUA) Malformation(s) not mentioned under brain, head/neck, chest, abdomen, extremities or spine - Code (a) |
| CUAH02b2 | CUA |  | 1 | 999 | 0.9987 | (CUA) Malformation(s) not mentioned under brain, head/neck, chest, abdomen, extremities or spine - Code (b) |
| CUBB01 | CUB |  | 1 | 999 | 0.0008 | (CUB) Were any structural abnormalities detected? |
| CUBB02a2 | CUB |  | 1 | 999 | 0.9417 | (CUB) Abnormality - Code (a) |
| CUBB02b2 | CUB |  | 1 | 999 | 0.9935 | (CUB) Abnormality - Code (b) |
| CUBB02c2 | CUB |  | 1 | 999 | 0.9974 | (CUB) Abnormality - Code (c) |
| CUBB02d2 | CUB |  | 1 | 999 | 0.9987 | (CUB) Abnormality - Code (d) |
| CUBB02e2 | CUB |  | 1 | 999 | 0.9992 | (CUB) Abnormality - Code (e) |
| CUBB02f2 | CUB |  | 1 | 999 | 0.9997 | (CUB) Abnormality - Code (f) |
| U1CB02 | U1C |  | 1 | ANY | 0.0014 | (U1C) Disclosable condition |
| U1CB04c | U1C |  | 1 | 999 | 0.9971 | (U1C) Disclosable condition reported - Incidental detection of complete or partial placenta previa or vasa previa |
| U1CC04 | U1C |  | 1 | MEAN | 0.0287 | (U1C) Left Uterine Artery - Peak Systolic Velocity (PSV) (cm/sec) |
| U1CC05 | U1C |  | 1 | MEAN | 0.0337 | (U1C) Left Uterine Artery - Systolic/Diastolic Ratio (S/D) |
| U1CC06 | U1C |  | 1 | MEAN | 0.0294 | (U1C) Left Uterine Artery - Resistance Index (RI) |
| U1CC07 | U1C |  | 1 | MEAN | 0.0309 | (U1C) Left Uterine Artery - Pulsatility Index (PI) |
| U1CC09 | U1C |  | 1 | 999 | 0.0266 | (U1C) Left Uterine Artery - Early diastolic notch present |
| U1CC12 | U1C |  | 1 | 999 | 0.023 | (U1C) Left Uterine Artery - Systolic notch present |
| U1CD04 | U1C |  | 1 | MEAN | 0.0345 | (U1C) Right Uterine Artery - Peak Systolic Velocity (PSV) (cm/sec) |
| U1CD05 | U1C |  | 1 | MEAN | 0.0402 | (U1C) Right Uterine Artery - Systolic/Diastolic Ratio (S/D) |
| U1CD06 | U1C |  | 1 | MEAN | 0.0345 | (U1C) Right Uterine Artery - Resistance Index (RI) |
| U1CD07 | U1C |  | 1 | MEAN | 0.0359 | (U1C) Right Uterine Artery - Pulsatility Index (PI) |
| U1CD09 | U1C |  | 1 | 999 | 0.0302 | (U1C) Right Uterine Artery - Early diastolic notch present |
| U1CD12 | U1C |  | 1 | 999 | 0.0294 | (U1C) Right Uterine Artery - Systolic notch present |
| U2AB02 | U2A |  | 2 | MEAN | 0.0113 | (U2A) Biparietal Diameter - cm |
| U2AB03 | U2A |  | 2 | MEAN | 0.011 | (U2A) Head Circumference - cm |
| U2AB04 | U2A |  | 2 | MEAN | 0.0115 | (U2A) Abdominal Circumference - cm |
| U2AB05 | U2A |  | 2 | MEAN | 0.0106 | (U2A) Femur Diaphysis Length - cm |
| U2AB06 | U2A |  | 2 | MEAN | 0.1167 | (U2A) Estimated fetal weight as reported - grams |
| U2AB07 | U2A |  | 2 | N/A | 0.595 | (U2A) Estimated fetal weight percentile as reported - percentile |
| U2AC01 | U2A |  | 2 | ANY | 0.4733 | (U2A) Disclosable condition |
| U2AC03c | U2A |  | 2 | 999 | 0.9871 | (U2A) Disclosable condition reported - Incidental finding of oligohydramnios |
| U2AC03e | U2A |  | 2 | 999 | 0.9871 | (U2A) Disclosable condition reported - Incidental detection of complete or partial placenta previa or vasa previa |
| U2BB02 | U2B |  | 2 | NEG1 | 0.0208 | (U2B) Cervical length - mm |
| U2BB03 | U2B |  | 2 | 999 | 0.0051 | (U2B) Funnel |
| U2BB04 | U2B |  | 2 | NEG1 | 0.9944 | (U2B) Funnel length - mm |
| U2BB05 | U2B |  | 2 | 999 | 0.0059 | (U2B) Debris |
| U2BC01 | U2B |  | 2 | ANY | 0.1082 | (U2B) Disclosable condition |
| U2BC03d | U2B |  | 2 | 999 | 0.9815 | (U2B) Disclosable condition reported - Incidental detection of complete or partial placenta previa or vasa previa |
| U2CB03 | U2C |  | 2 | ANY | 0 | (U2C) Disclosable condition reported |
| U2CB04c | U2C |  | 2 | 999 | 0.9825 | (U2C) Disclosable condition reported - Incidental detection of complete or partial placenta previa or vasa previa |
| U2CC04 | U2C |  | 2 | MEAN | 0.0167 | (U2C) Left Uterine Artery - Peak Systolic Velocity (PSV) (cm/sec) |
| U2CC05 | U2C |  | 2 | MEAN | 0.0191 | (U2C) Left Uterine Artery - Systolic/Diastolic Ratio (S/D) |
| U2CC06 | U2C |  | 2 | MEAN | 0.0171 | (U2C) Left Uterine Artery - Resistance Index (RI) |
| U2CC07 | U2C |  | 2 | MEAN | 0.0188 | (U2C) Left Uterine Artery - Pulsatility Index (PI) |
| U2CC09 | U2C |  | 2 | 999 | 0.0146 | (U2C) Left Uterine Artery - Early diastolic notch present |
| U2CC12 | U2C |  | 2 | 999 | 0.016 | (U2C) Left Uterine Artery - Systolic notch present |
| U2CD04 | U2C |  | 2 | MEAN | 0.0151 | (U2C) Right Uterine Artery - Peak Systolic Velocity (PSV) (cm/sec) |
| U2CD05 | U2C |  | 2 | MEAN | 0.0182 | (U2C) Right Uterine Artery - Systolic/Diastolic Ratio (S/D) |
| U2CD06 | U2C |  | 2 | MEAN | 0.0151 | (U2C) Right Uterine Artery - Resistance Index (RI) |
| U2CD07 | U2C |  | 2 | MEAN | 0.0164 | (U2C) Right Uterine Artery - Pulsatility Index (PI) |
| U2CD09 | U2C |  | 2 | 999 | 0.0131 | (U2C) Right Uterine Artery - Early diastolic notch present |
| U2CD12 | U2C |  | 2 | 999 | 0.0148 | (U2C) Right Uterine Artery - Systolic notch present |
| U3AB02 | U3A |  | 3 | MEAN | 0.0077 | (U3A) Biparietal Diameter (cm) |
| U3AB03 | U3A |  | 3 | MEAN | 0.0076 | (U3A) Head Circumference (cm) |
| U3AB04 | U3A |  | 3 | MEAN | 0.0064 | (U3A) Abdominal Circumference (cm) |
| U3AB05 | U3A |  | 3 | MEAN | 0.0068 | (U3A) Femur Diaphysis Length (cm) |
| U3AB07 | U3A |  | 3 | MEAN | 0.0127 | (U3A) Estimated fetal weight as reported - grams |
| U3AC01 | U3A |  | 3 | ANY | 0.0437 | (U3A) Disclosable condition |
| U3AC03c | U3A |  | 3 | SUM.LEQ | 0.9915 | (U3A) Disclosable condition reported - Oligohydramnios (maximal vertical pocket of <2 cm or AFI <5 cm) |
| U3AC03e | U3A |  | 3 | 999 | 0.9915 | (U3A) Disclosable condition reported - Incidental detection of complete or partial placenta previa or vasa previa |
| U3BB02 | U3B |  | 3 | NEG1 | 0.0152 | (U3B) Cervical length - mm |
| U3BB03 | U3B |  | 3 | 999 | 0.0052 | (U3B) Funnel |
| U3BB04 | U3B |  | 3 | NEG1 | 0.9799 | (U3B) Funnel length - mm |
| U3BB05 | U3B |  | 3 | 999 | 0.0064 | (U3B) Debris |
| U3BC01 | U3B |  | 3 | ANY | 0.0122 | (U3B) Disclosable condition |
| U3BC03d | U3B |  | 3 | 999 | 0.984 | (U3B) Disclosable condition reported - Incidental detection of complete or partial placenta previa or vasa previa |
| U3CB02 | U3C |  | 3 | ANY | 0.0009 | (U3C) Disclosable condition |
| U3CB04c | U3C |  | 3 | 999 | 0.9954 | (U3C) Disclosable condition reported - Incidental detection of complete or partial placenta previa or vasa previa |
| U3CC04 | U3C |  | 3 | MEAN | 0.0271 | (U3C) Left Uterine Artery - Peak Systolic Velocity (PSV) (cm/sec) |
| U3CC05 | U3C |  | 3 | MEAN | 0.0271 | (U3C) Left Uterine Artery - Systolic/Diastolic Ratio (S/D) |
| U3CC06 | U3C |  | 3 | MEAN | 0.0264 | (U3C) Left Uterine Artery - Resistance Index (RI) |
| U3CC07 | U3C |  | 3 | MEAN | 0.0266 | (U3C) Left Uterine Artery - Pulsatility Index (PI) |
| U3CC09 | U3C |  | 3 | 999 | 0.0243 | (U3C) Left Uterine Artery - Early diastolic notch present |
| U3CC12 | U3C |  | 3 | 999 | 0.0254 | (U3C) Left Uterine Artery - Systolic notch present |
| U3CD04 | U3C |  | 3 | MEAN | 0.0259 | (U3C) Right Uterine Artery - Peak Systolic Velocity (PSV) (cm/sec) |
| U3CD05 | U3C |  | 3 | MEAN | 0.0275 | (U3C) Right Uterine Artery - Systolic/Diastolic Ratio (S/D) |
| U3CD06 | U3C |  | 3 | MEAN | 0.0257 | (U3C) Right Uterine Artery - Resistance Index (RI) |
| U3CD07 | U3C |  | 3 | MEAN | 0.0264 | (U3C) Right Uterine Artery - Pulsatility Index (PI) |
| U3CD09 | U3C |  | 3 | 999 | 0.0234 | (U3C) Right Uterine Artery - Early diastolic notch present |
| U3CD12 | U3C |  | 3 | 999 | 0.0268 | (U3C) Right Uterine Artery - Systolic notch present |
| U3DC01 | U3D |  | 3 | 999 | 0.0677 | (U3D) Fetal adrenal gland mass |
| U3DC02 | U3D |  | 3 | 999 | 0.0004 | (U3D) Disclosable condition reported |
| U3DD01 | U3D |  | 3 | 999 | 0.0895 | (U3D) Fetal Adrenal Gland Measures Visit 3 - Which gland was used for measurement |
| Calc_Polyhydramnios |  | Calc_Polyhydramnios | 3 |  |  |  |
| U3AB06a | U3A |  | 3 | MEAN | 0.0761 | (U3A) Amniotic Fluid Index (AFI) - Quadrant 1 (cm) |
| U3AB06b | U3A |  | 3 | MEAN | 0.074 | (U3A) Amniotic Fluid Index (AFI) - Quadrant 2 (cm) |
| U3AB06c | U3A |  | 3 | MEAN | 0.0736 | (U3A) Amniotic Fluid Index (AFI) - Quadrant 3 (cm) |
| U3AB06d | U3A |  | 3 | MEAN | 0.0833 | (U3A) Amniotic Fluid Index (AFI) - Quadrant 4 (cm) |
| adrenal_gland_length |  | adrenal_measure | 3 |  |  |  |
| U3DD02a1 | U3D |  | 3 | MEAN | 0.1898 | (U3D) Adrenal Gland Measurements, Adrenal gland - Length in mm (1) |
| U3DD02a2 | U3D |  | 3 | MEAN | 0.2111 | (U3D) Adrenal Gland Measurements, Adrenal gland - Length in mm (2) |
| U3DD02a3 | U3D |  | 3 | MEAN | 0.257 | (U3D) Adrenal Gland Measurements, Adrenal gland - Length in mm (3) |
| U3DD02a1 | U3D |  | 3 | MEAN | 0.9951 | (U3D) Repeat Adrenal Gland Measurements, Adrenal gland length in mm, Measure 1 |
| U3DF02a2 | U3D |  | 3 | MEAN | 0.9955 | (U3D) Repeat Adrenal Gland Measurements, Adrenal gland length in mm, Measure 2 |
| U3DF02a3 | U3D |  | 3 | MEAN | 0.9955 | (U3D) Repeat Adrenal Gland Measurements, Adrenal gland length in mm, Measure 3 |
| U3DD05 | U3D |  | 3 | N/A | 0.0579 | (U3D) Fetal Adrenal Gland Measures Visit 3 - Adrenal Glad measurements exam complete |
| U3DF05 | U3D |  | 3 | N/A | 0.9933 | (U3D) Repeat Adrenal Gland Measurements - Exam complete |
| **fetal_zone_length** |  | adrenal_measure | 3 |  |  |  |
| U3DD02b1 | U3D |  | 3 | MEAN | 0.1906 | (U3D) Adrenal Gland Measurements, Fetal zone - Length in mm (1) |
| U3DD02b2 | U3D |  | 3 | MEAN | 0.212 | (U3D) Adrenal Gland Measurements, Fetal zone - Length in mm (2) |
| U3DD02b3 | U3D |  | 3 | MEAN | 0.2584 | (U3D) Adrenal Gland Measurements, Fetal zone - Length in mm (3) |
| U3DF02b1 | U3D |  | 3 | MEAN | 0.9951 | (U3D) Repeat Adrenal Gland Measurements, Fetal zone length in mm, Measure 1 |
| U3DF02b2 | U3D |  | 3 | MEAN | 0.9955 | (U3D) Repeat Adrenal Gland Measurements, Fetal zone length in mm, Measure 2 |
| U3DF02b3 | U3D |  | 3 | MEAN | 0.9955 | (U3D) Repeat Adrenal Gland Measurements, Fetal zone length in mm, Measure 3 |
| U3DD05 | U3D |  | 3 | N/A | 0.0579 | (U3D) Fetal Adrenal Gland Measures Visit 3 - Adrenal Glad measurements exam complete |
| U3DF05 | U3D |  | 3 | N/A | 0.9933 | (U3D) Repeat Adrenal Gland Measurements - Exam complete |
| **adrenal_gland_width** |  | adrenal_measure | 3 |  |  |  |
| U3DD03a1 | U3D |  | 3 | MEAN | 0.1929 | (U3D) Adrenal Gland Measurements, Adrenal gland - Width in mm (1) |
| U3DD03a2 | U3D |  | 3 | MEAN | 0.2134 | (U3D) Adrenal Gland Measurements, Adrenal gland - Width in mm (2) |
| U3DD03a3 | U3D |  | 3 | MEAN | 0.2615 | (U3D) Adrenal Gland Measurements, Adrenal gland - Width in mm (3) |
| U3DF03a1 | U3D |  | 3 | MEAN | 0.9951 | (U3D) Repeat Adrenal Gland Measurements, Adrenal gland length in mm, Measure 1 |
| U3DF03a2 | U3D |  | 3 | MEAN | 0.9955 | (U3D) Repeat Adrenal Gland Measurements, Adrenal gland length in mm, Measure 2 |
| U3DF03a3 | U3D |  | 3 | MEAN | 0.9955 | (U3D) Repeat Adrenal Gland Measurements, Adrenal gland length in mm, Measure 3 |
| U3DD05 | U3D |  | 3 | N/A | 0.0579 | (U3D) Fetal Adrenal Gland Measures Visit 3 - Adrenal Glad measurements exam complete |
| U3DF05 | U3D |  | 3 | N/A | 0.9933 | (U3D) Repeat Adrenal Gland Measurements - Exam complete |
| **fetal_zone_width** |  | adrenal_measure | 3 |  |  |  |
| U3DD03b1 | U3D |  | 3 | MEAN | 0.1929 | (U3D) Adrenal Gland Measurements, Fetal zone - Width in mm (1) |
| U3DD03b2 | U3D |  | 3 | MEAN | 0.2143 | (U3D) Adrenal Gland Measurements, Fetal zone - Width in mm (2) |
| U3DD03b3 | U3D |  | 3 | MEAN | 0.2633 | (U3D) Adrenal Gland Measurements, Fetal zone - Width in mm (3) |
| U3DF03b1 | U3D |  | 3 | MEAN | 0.9951 | (U3D) Repeat Adrenal Gland Measurements, Fetal zone width in mm, Measure 1 |
| U3DF03b2 | U3D |  | 3 | MEAN | 0.9955 | (U3D) Repeat Adrenal Gland Measurements, Fetal zone width in mm, Measure 2 |
| U3DF03b3 | U3D |  | 3 | MEAN | 0.9955 | (U3D) Repeat Adrenal Gland Measurements, Fetal zone width in mm, Measure 3 |
| U3DD05 | U3D |  | 3 | N/A | 0.0579 | (U3D) Fetal Adrenal Gland Measures Visit 3 - Adrenal Glad measurements exam complete |
| U3DF05 | U3D |  | 3 | N/A | 0.9933 | (U3D) Repeat Adrenal Gland Measurements - Exam complete |
| **adrenal_gland_depth** |  | adrenal_measure | 3 |  |  |  |
| U3DD04a1 | U3D |  | 3 | MEAN | 0.7657 | (U3D) Adrenal Gland Measurements, Adrenal gland - Depth in mm (1) |
| U3DD04a2 | U3D |  | 3 | MEAN | 0.7679 | (U3D) Adrenal Gland Measurements, Adrenal gland - Depth in mm (2) |
| U3DD04a3 | U3D |  | 3 | MEAN | 0.7719 | (U3D) Adrenal Gland Measurements, Adrenal gland - Depth in mm (3) |
| U3DF04a1 | U3D |  | 3 | MEAN | 0.9982 | (U3D) Repeat Adrenal Gland Measurements, Adrenal gland depth in mm, Measure 1 |
| U3DF04a2 | U3D |  | 3 | MEAN | 0.9982 | (U3D) Repeat Adrenal Gland Measurements, Adrenal gland depth in mm, Measure 2 |
| U3DF04a3 | U3D |  | 3 | MEAN | 0.9982 | (U3D) Repeat Adrenal Gland Measurements, Adrenal gland depth in mm, Measure 3 |
| U3DD05 | U3D |  | 3 | N/A | 0.0579 | (U3D) Fetal Adrenal Gland Measures Visit 3 - Adrenal Glad measurements exam complete |
| U3DF05 | U3D |  | 3 | N/A | 0.9933 | (U3D) Repeat Adrenal Gland Measurements - Exam complete |
| **fetal_zone_depth** |  | adrenal_measure | 3 |  |  |  |
| U3DD04b1 | U3D |  | 3 | MEAN | 0.7657 | (U3D) Adrenal Gland Measurements, Fetal zone - Depth in mm (1) |
| U3DD04b2 | U3D |  | 3 | MEAN | 0.7679 | (U3D) Adrenal Gland Measurements, Fetal zone - Depth in mm (2) |
| U3DD04b3 | U3D |  | 3 | MEAN | 0.7719 | (U3D) Adrenal Gland Measurements, Fetal zone - Depth in mm (3) |
| U3DF04b1 | U3D |  | 3 | MEAN | 0.9982 | (U3D) Repeat Adrenal Gland Measurements, Fetal zone, depth in mm, Measure 1 |
| U3DF04b2 | U3D |  | 3 | MEAN | 0.9982 | (U3D) Repeat Adrenal Gland Measurements, Fetal zone, depth in mm, Measure 2 |
| U3DF04b3 | U3D |  | 3 | MEAN | 0.9982 | (U3D) Repeat Adrenal Gland Measurements, Fetal zone, depth in mm, Measure 3 |
| U3DD05 | U3D |  | 3 | N/A | 0.0579 | (U3D) Fetal Adrenal Gland Measures Visit 3 - Adrenal Glad measurements exam complete |
| U3DF05 | U3D |  | 3 | N/A | 0.9933 | (U3D) Repeat Adrenal Gland Measurements - Exam complete |

### 6.8 Medical History

The medical history filtering includes questions regarding the current or recent (prenatal labs) medical conditions of the patient, which is a quite large category. As a result, using literature review and clinician input, we narrowed down our relevant features to PTB-related factors like STI screens, gynecological infections, and diabetes/hypertension data. Aside from diagnostics, we also included prior information about previous pregnancy loss history, given the trial’s focus on nulliparous patients, as well as relevant CBC results like hematocrit levels, and patient previous procedures and pre-pregnancy weight.

**Total Features:** 2294

**Layer 1 Features:** 100

**Layer 2 Features:** 100

**Relevant Files:** CLA, CLB, CMA, CMB, CMD, CME, S02, V1A, VXX, demographics (ancillary), pregnancy outcomes (ancillary)

#### Dropped Features

For this group, we dropped most features due to a large amount of information. As a result, we focused on health conditions that pertained most to PTB risk factors.

**Table 9:**
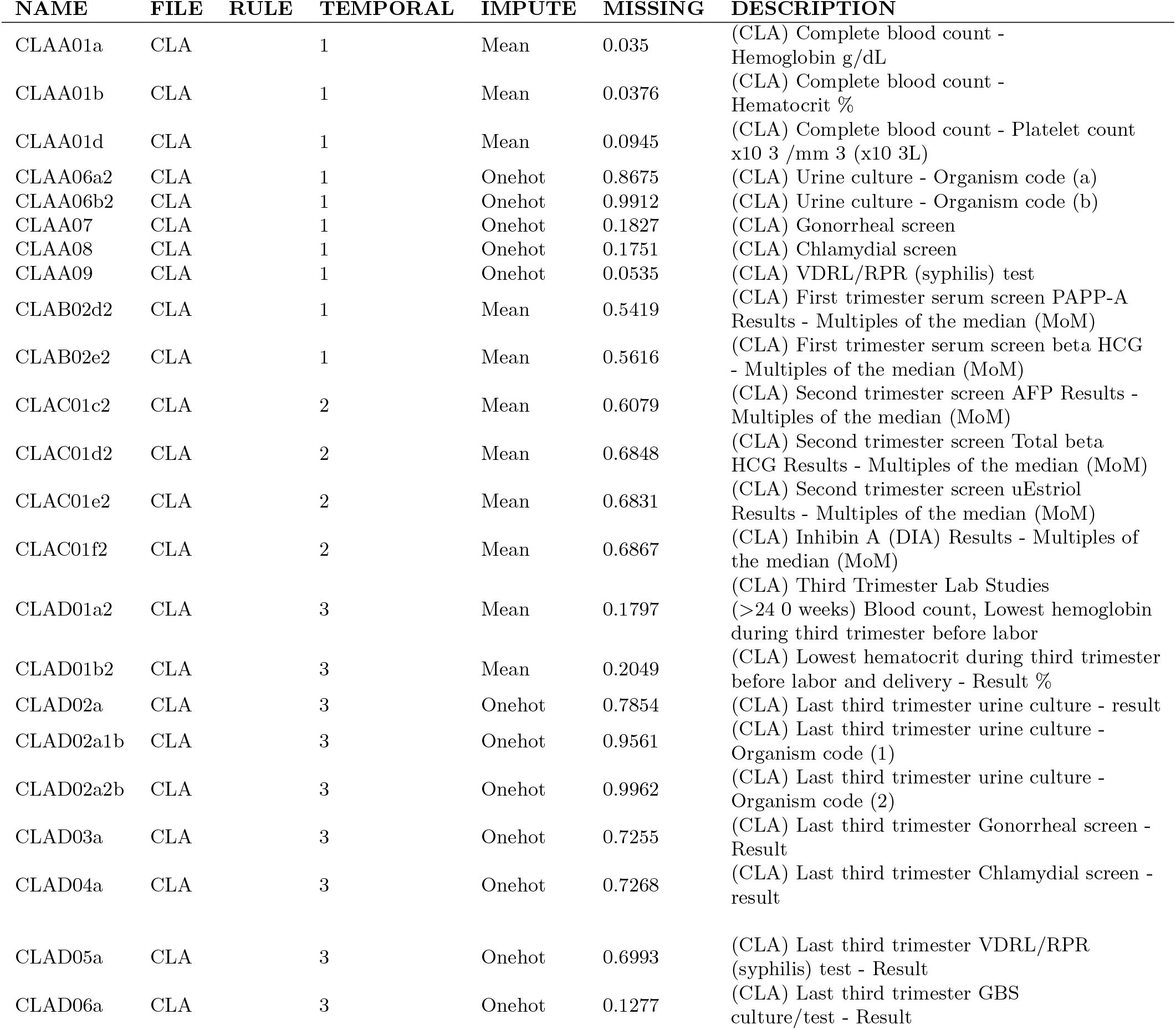
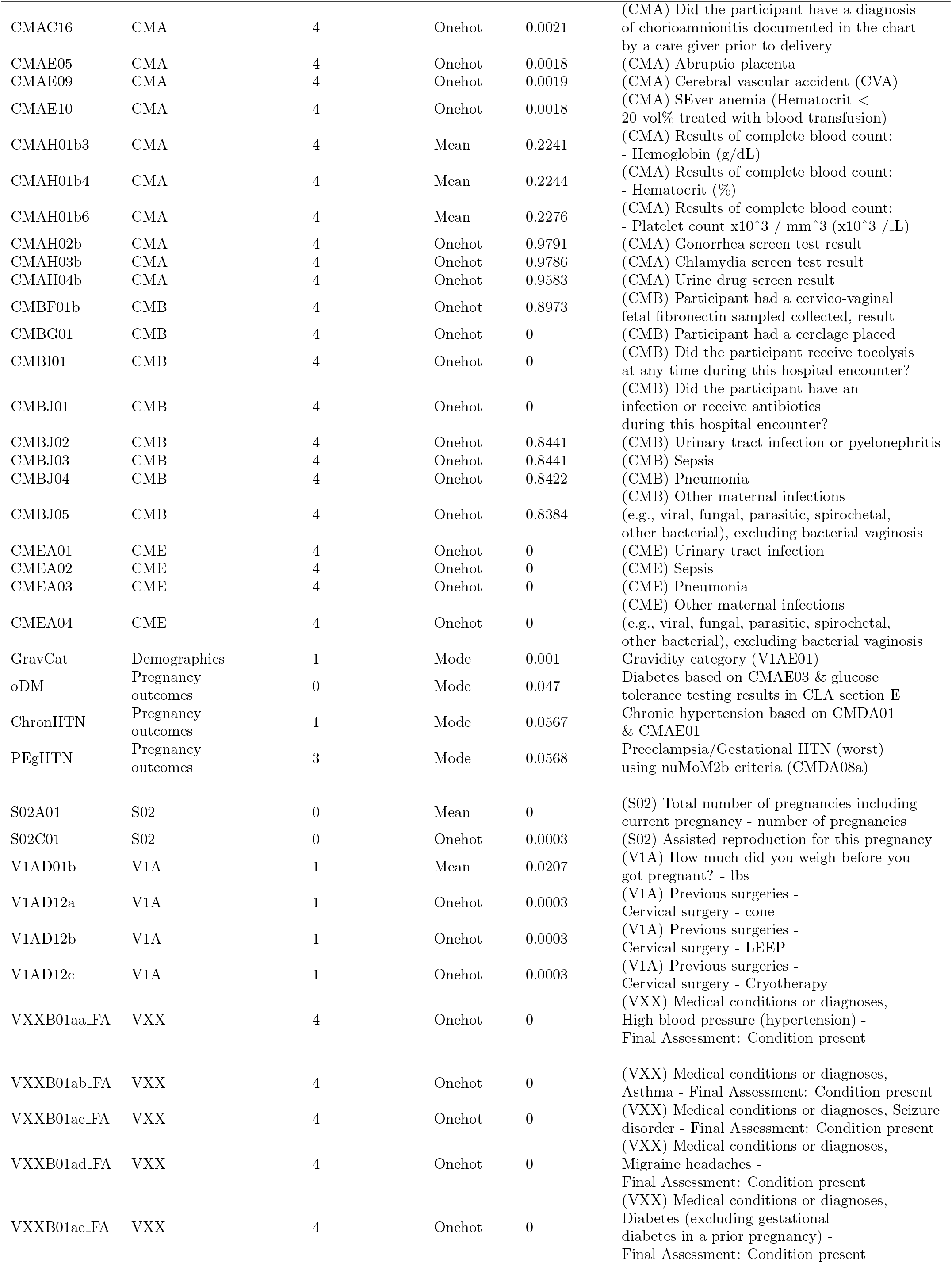
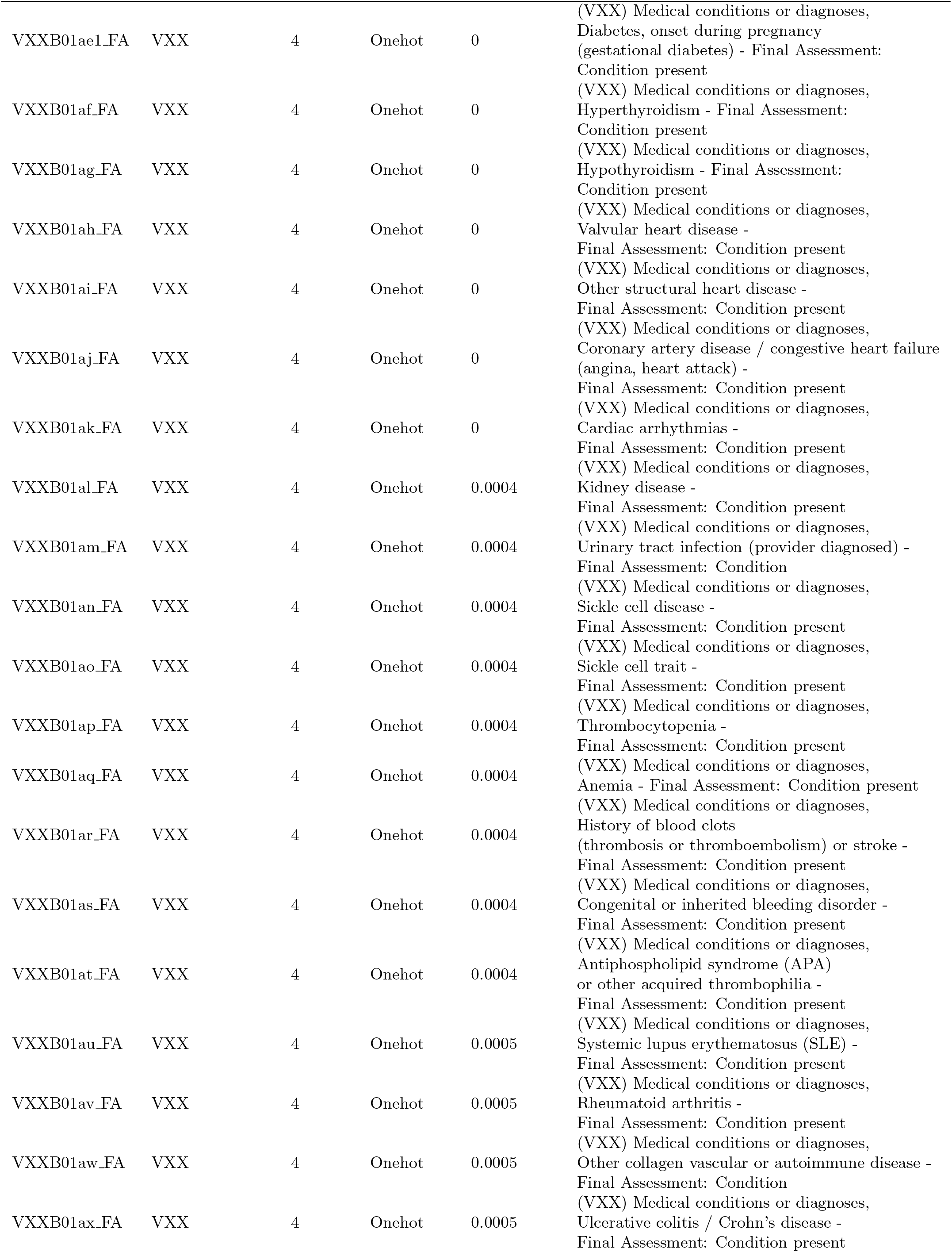
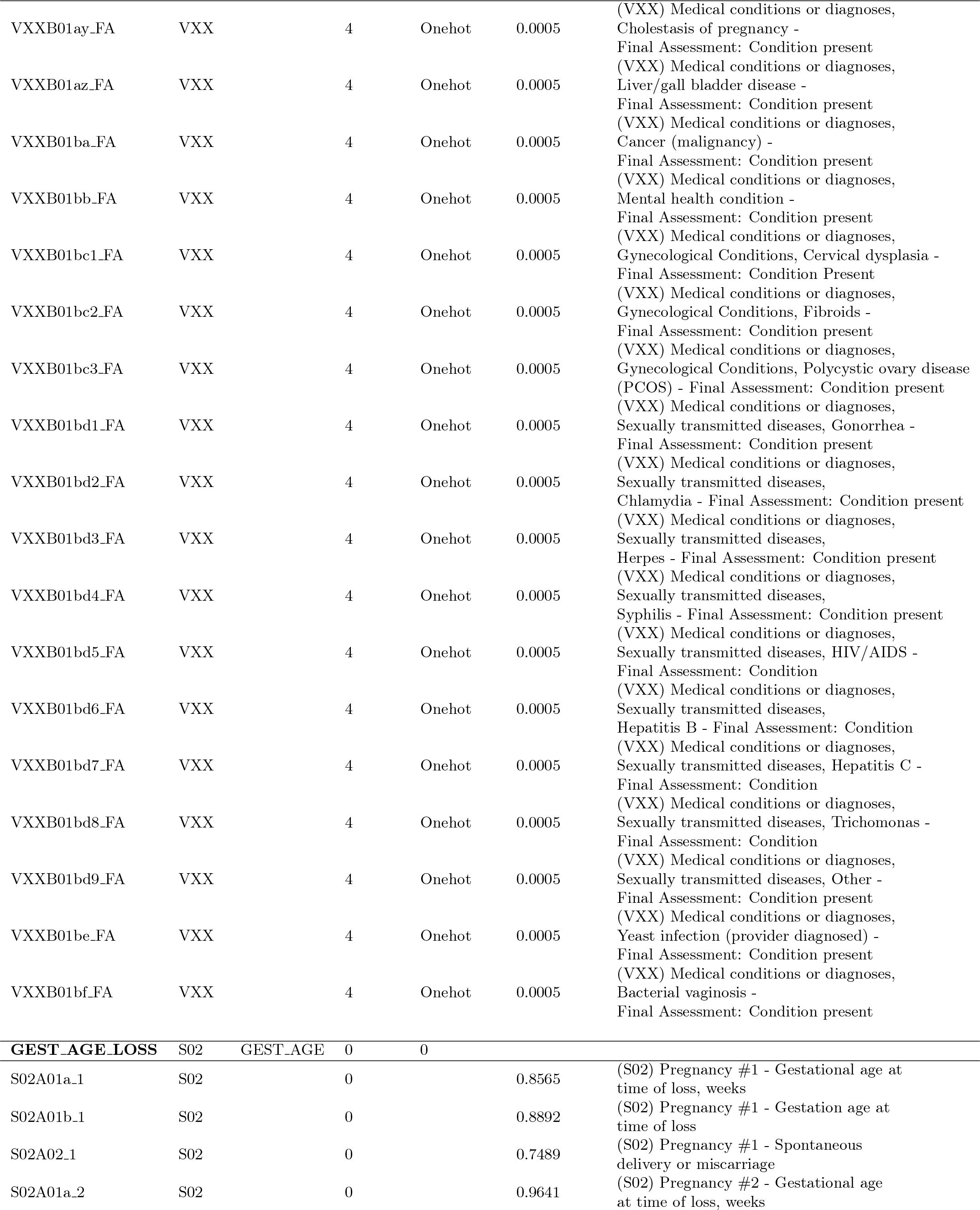
Medical History Filtering

### 6.9 Outcomes

The outcomes filtering concerns post-pregnancy analysis of the newborn (all kinds of delivery), and variables that are highly predictive of preterm birth, such as major fetal conditions. Some of these variables may be used as class labels, such as pOUTCOME or GAwksEND, and others may provide privileged information for certain models. **The vast majority of this data is highly prone to class leakage and should not be used in modeling, but is used for data analysis**. Provided in the table are a **sample** of features that are useful in data analysis or as class labels.

**Total # Features:** 1165

**Relevant Files:** CBA, CBB, CBC, CMA, CMC, CPA, pregnancy outcomes, S02, U02, U2A, U3A, V4A

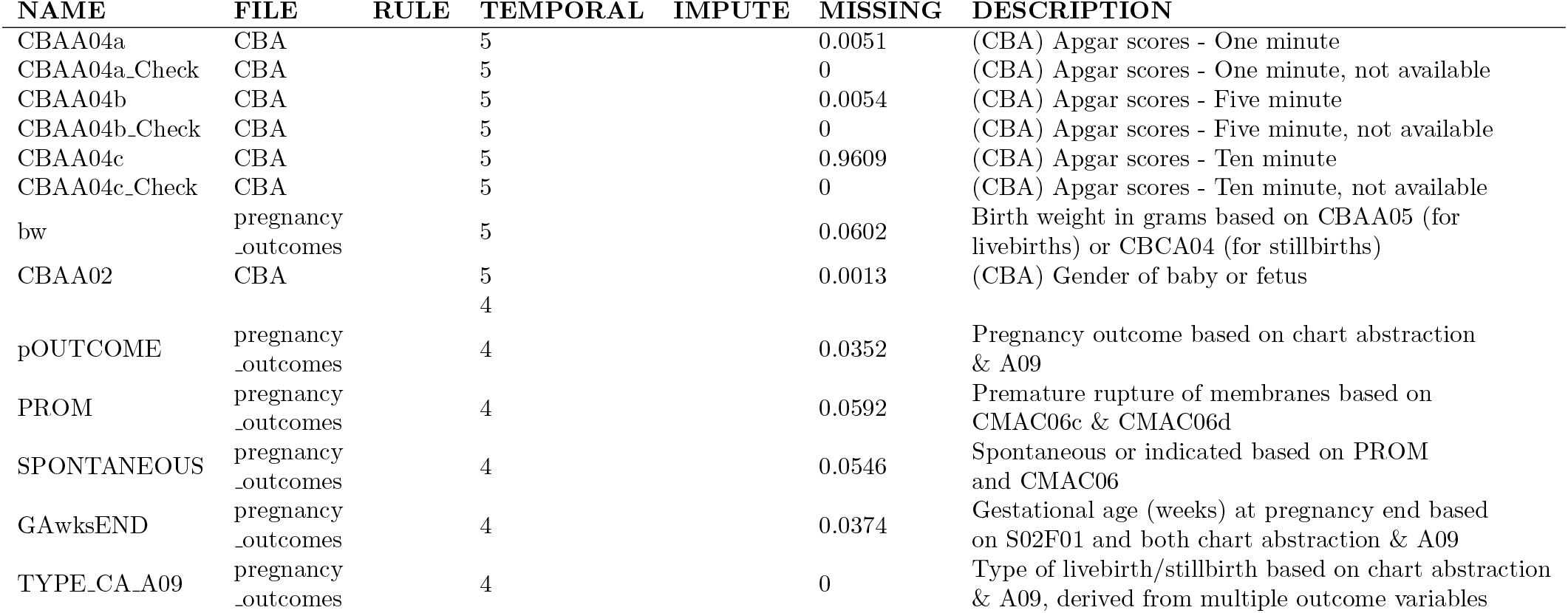

### 6.10 Treatment

The treatment filtering contains features that pertain to drug administration directly related to delivery or the prevention of preterm birth. This includes administration of steroids for fetal lung maturation, tocolytics, progesterone, and various classes of drugs used specifically for delivery. **These features are highly predictive of PTB, and are thus only used for analysis or sequential treatment decision making. Shown is a sample of these features**.

**Total # Features:** 149

**Layer 1 # Features:** N/A (27 Available)

**Layer 2 # Features:** N/A (34 Available)

**Relevant Files:** CMA

#### Dropped Features

For this group, there was much metadata that was used to place the administration within a particular time point, and then later dropped. Medication codes were used instead of names. Details on which particular drug within a class was used were dropped in L1, as the intended and possible effect were known and similar. Anticonvulsant details were returned in L2 as they can have dramatic differences.

| NAME | FILE | RULE | TEMPORAL | IMPUTE | MISSINGNESS | DESCRIPTION |
| --- | --- | --- | --- | --- | --- | --- |
| CMAF01 | CMA |  | Date Abstraction | ANY | 0.002 | (CMA) Did the Participant receive steroids for fetal lung maturation at any time during this pregnancy? |
| CMAF02 | CMA |  | Date Abstraction | ANY | 0.0023 | (CMA) Did the Participant receive progesterone at any time during this pregnancy? |
| CMAF01a | CMA |  | Date Abstraction | ANY | 0.0011 | (CMA) Steroids for fetal lung maturation: Betamethasone |
| CMAF01a3 | CMA |  | Date Abstraction | ANY | 0.9544 | (CMA) Steroids for fetal lung maturation: Betamethasone - Total number of injections given over pregnancy |
| CMAF01b | CMA |  | Date Abstraction | ANY | 0.0011 | (CMA) Steroids for fetal lung maturation: Dexamethasone - Dexamethasone |
| CMAF01b3 | CMA |  | Date Abstraction | ANY | 0.9992 | (CMA) Steroids for fetal lung maturation: Dexamethasone - Total number of doses given over pregnancy |
| CMAF02a | CMA |  | Date Abstraction | 999 | 0.981 | (CMA) Progesterone vehicle of administration: |
| CMAF02a_sp | CMA |  | Date Abstraction | 999 | 0.9979 | (CMA) Progesterone vehicle of administration: - Other, specify |
| CMAF02b1 | CMA |  | Date Abstraction | -1 | 0.9979 | (CMA) Progesterone dose - % |
| CMAF02b2 | CMA |  | Date Abstraction | -1 | 0.989 | (CMA) Progesterone dose - mg |
| CMAF02c | CMA |  | Date Abstraction | 0 | 0.9743 | (CMA) Progesterone frequency of administration |
| CMAF02c_sp | CMA |  | Date Abstraction | 999 | 0.9875 | (CMA) Progesterone frequency of administration - Other, specify |
| CMAG01 | CMA |  | 4 | ANY | 0.0021 | (CMA) Did the Participant receive tocolysis at any time during this hospitalization for delivery? |
| CMAG02 | CMA |  | 4 | ANY | 0.0021 | (CMA) Were antibiotics used anytime during this hospitalization for delivery? |
| CMAG03 | CMA |  | 4 | ANY | 0.002 | (CMA) Were antihypertensives used anytime during this hospitalization for delivery? |
| CMAG04 | CMA |  | 4 | ANY | 0.002 | (CMA) Were anticonvulsants used anytime during this hospitalization for delivery? |
| CMAG05 | CMA |  | 4 | ANY | 0.0018 | (CMA) Was magnesium sulfate given specifically for neuroprophylaxis anytime during this hospitalization for delivery? |
| CMAG06 | CMA |  | 4 | ANY | 0.0021 | (CMA) Were any other prescribed medications used during this hospitalization for delivery? |
| CMAG06a | CMA |  | 4 | 2 | 0.0011 | (CMA) Other prescribed medications used during this hospitalization for delivery - Insulin |
| CMAG06b | CMA |  | 4 | 2 | 0.0011 | (CMA) Other prescribed medications used during this hospitalization for delivery - Heparin / LMW heparin injection |
| CMAG06c1 | CMA |  | 4 | 777 | 0.7016 | (CMA) Other prescribed medications used during this hospitalization for delivery - Medication code (1) |
| CMAG06d1 | CMA |  | 4 | 777 | 0.871 | (CMA) Other prescribed medications used during this hospitalization for delivery - Medication code (2) |
| CMAG06e1 | CMA |  | 4 | 777 | 0.9373 | (CMA) Other prescribed medications used during this hospitalization for delivery - Medication code (3) |
| CMAG06f1 | CMA |  | 4 | 777 | 0.9667 | (CMA) Other prescribed medications used during this hospitalization for delivery - Medication code (4) |
| CMAG06g1 | CMA |  | 4 | 777 | 0.9821 | (CMA) Other prescribed medications used during this hospitalization for delivery - Medication code (5) |
| CMAG06h1 | CMA |  | 4 | 777 | 0.9888 | (CMA) Other prescribed medications used during this hospitalization for delivery - Medication code (6) |
| CMAG06i1 | CMA |  | 4 | 777 | 0.9935 | (CMA) Other prescribed medications used during this hospitalization for delivery - Medication code (7) |

### 6.11 Food Frequency Analysis

The food frequency analysis file takes into account the food and nutrients consumed by patients in the 3 months prior to conception. We were most interested in the calculated nutrient intake as opposed to less interpretable data like grams of food consumed or food pyramid group quantities. In addition to nutrients, we also attempted to capture the overall energy intake by using glycemic load and calorie intake as proxies.

**Total Features:** 737

**Layer 1 Features:** 38

**Layer 2 Features:** 38

**Relevant Files:** food frequency analysis (ancillary)

#### Dropped Features

For this group, we dropped the features that contained redundant information about food intake that was already captured by the vitamin amounts. For instance, quantities of food items such as “glasses of milk” would be reflected in the overall vitamin and calorie consumption.

**Table 10:**
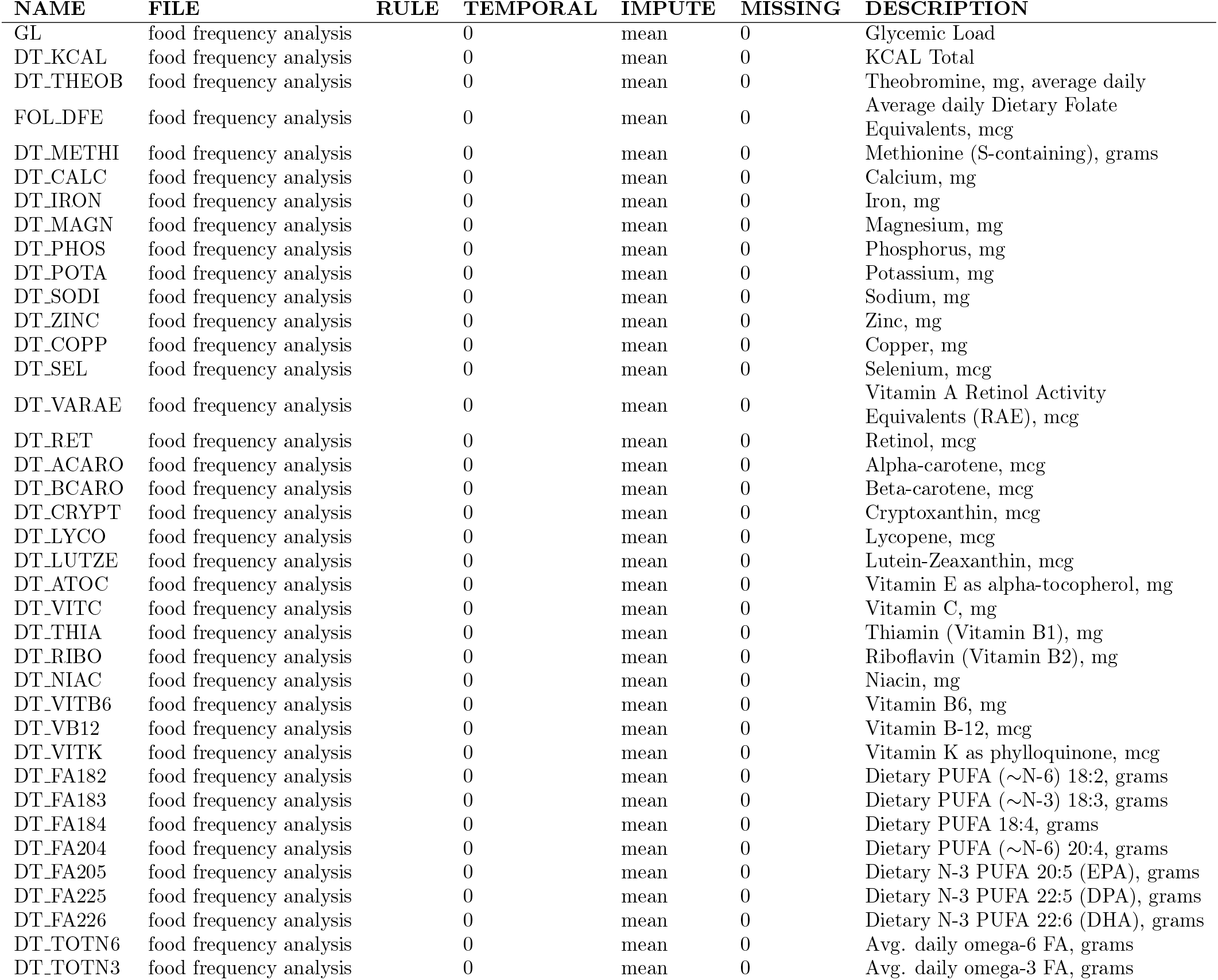
Food Frequency Filtering

### 6.12 Sleep Substudy

The sleep substudy filtering includes sleep quantity and quality by calculating the average hours slept per night as well as sleep apnea diagnoses.

**Total Features:** 6

**Layer 1 Features:** 4

**Layer 2 Features:** 6

**Relevant Files:** V1L, V3L

#### Dropped Features

Due to sleep being a separate substudy, only a few features were selected to be included. The rest were dropped as of writing.

**Table 11:** Sleep Substudy Filtering

| NAME | FILE | RULE | TEMPORAL | IMPUTE | MISSING | DESCRIPTION |
| --- | --- | --- | --- | --- | --- | --- |
| V1LF01a | V1L |  | 1 | Onehot | 0.00514 | Have you ever been told by a doctor or other health professionals that you have sleep apnea or obstructive sleep apnea? |
| V3LF01a | V3L |  | 3 | Onehot | 0.00464 | Have you ever been told by a doctor or other health professionals that you have sleep apnea or obstructive sleep apnea? |
| **V1A_SLEEP** | V1L | SLEEP_AVG | 1 | Mean |  |  |
| V1LA02a | V1L |  | 1 |  | 0.00463 | How many hours of sleep do you usually get per night: On weekdays or workdays? |
| V1LA02b | V1L |  | 1 |  | 0.00463 | How many hours of sleep do you usually get per night: On weekends? |
| **V3A_SLEEP** | V3L | SLEEP_AVG | 3 | Mean |  |  |
| V3LA02a | V3L |  | 3 |  | 0.00357 | How many hours of sleep do you usually get per night: On weekdays or workdays? |
| V3LA02b | V3L |  | 3 |  | 0.00357 | How many hours of sleep do you usually get per night: On weekends? |

### 6.13 Genetic data

The genotyped cohort comprises 9,757 nulliparous women from the nuMoM2b study who had adequate samples and agreed to be genotyped. DNA extractions from whole blood, which had been frozen at −80°, were carried out on a Qiasymphony instrument at the Center for Bioinformatics and Genomics (Indiana University). Genotyping was done at the Van Andel Institute (Grand Rapids, MI, USA) using the Infinium Multi-Ethnic Global D2 BeadChip (Illumina, Miami, USA). We imposed standard filters for quality control of loci at this stage (cluster separation < 0.3, AA R Mean < 0.2, AB R Mean < 0.2, BB R Mean < 0.2, 10% GC < 0.3) using GenomeStudio v2.4 (Illumina). Genotype calls (in .GCT format) for the 1,748,280 loci that passed initial quality control were made with Beeline autoconvert (Ilumina).

## 7 Lessons Learned and Discussion

The main goals of the data preprocessing of the nuMoM2b dataset were to get a deeper understanding of the data, extract the most relevant features, and reduce the dimensionality of the data. Additionally, we wanted to make the fewest assumptions when filling in unknown values, and to understand the intricacies behind the dependencies present in the data.

The original ratio of features to patients of around 4,600 to 10,000 is too high to create reliable models because of the high possibility of over-fitting in a high dimensional space. Therefore, it was crucial for the interdisciplinary team to work together to leverage medical expertise, computing and statistics skills in order to gain a good grasp of the wealth of information in the nuMoM2b dataset. The data preprocessing team worked to make sure that any medical assumptions, such as grouping related conditions together or deciding to drop select details, were approved by the medical experts.

It was equally crucial to understand the protocol that doctors follow in the administration of interventions. Debates about definitions, classification of conditions, and treatments exist and were taken into consideration. A notable example is the debate around the usage of progesterone as an intervention for preterm birth. Another is simply the definitions of spontaneous and indicated preterm birth and how the events that fall under those categories may have changed over time.

With regards to the data itself, the most significant challenges were understanding the dependencies between different features and their medical relevance to preterm birth. The data includes both patient interviews and abstractions from the patients’ charts. Sometimes the questions overlapped, other times they were parallel, or had features that combined information from both. Each source has a different level of relevance in building the models.

Standardization of data formatting was another obstacle. Much of the original labeling and information provided in the codebooks was not sufficient to directly begin the modeling process. For instance, labels such as variable type had to be manually added. It was also important to determine when specific data were relevant and when they were collected. For example, for data that were not collected exactly within the strict time designations for Visits 1 through 4, the associated dates were computed relative to the estimated date for the start of pregnancy. Manual inspection was also required when coded values of responses did not match the information shown in the data collection forms. Codings such as *Don’t Know*, or *Refused response* did not exist in the data even though they were mentioned in the codebooks.

Throughout the preprocessing, we aimed to make the filtering and imputation steps as systematic as possible, while organizing the data into medically homogeneous groups. The discussion here is merely a glimpse of the intended goals and actions taken during the preprocessing. As our team delves into the PTB-related research goals, we anticipate that more preprocessing will be required on specific data.

## 8 Institutional review board statement and funding sources

Human subjects approval for this study, titled “SCH: Prediction of Preterm Birth in Nulliparous Women”, was obtained following review by Columbia University Human Subjects Institutional Review Board under number IRB-AAAR9413, and the City University of New York CUNY HRPP/IRB review number 2019-0855. Human subjects training requirements were completed by all authors of this study.

This work is supported by NIH/NLM (www.nlm.nih.gov) grant R01LM013327.

## Data Availability

The dataset is now available through Dash, upon request

https://www.nichd.nih.gov/research/supported/nuMoM2b

## Notes

### Competing Interest Statement

The authors have declared no competing interest.

### Funding Statement

This work was supported by NIH/NLM (www.nlm.nih.gov) grant R01LM013327

### Author Declarations

Human subjects approval for this study, titled "SCH: Prediction of Preterm Birth in Nulliparous Women", was obtained following review by Columbia University Human Subjects Institutional Review Board under number IRB-AAAR9413, and the City University of New York CUNY HRPP/IRB review number 2019-0855.

